# Plasma proteomic signatures of social support and their association with cardiovascular disease and mortality

**DOI:** 10.1101/2025.08.07.25333199

**Authors:** Pei Qin, Jessica Gong, Andrew Steptoe, Daisy Fancourt

## Abstract

**Background:** Social support has been related to cardiovascular disease (CVD) incidence and mortality in longitudinal cohort analyses, but the biological pathways underpinning this remain underexplored. We explored the protein signatures of social support amongst older adults and the mediating effect of proteins in the association between social support and CVD and mortality.

**Methods:** Data from 3,141 adults over the age of 50 in the English Longitudinal Study of Ageing who had plasma proteome data were analysed, with CVD and mortality outcomes followed up for 16 years following through Hospital Episode Statistics and the National Health Service Central Registry. Linear and Cox regression analyses were used to identify proteins associated with social support, CVD and mortality. Mediation analysis was then performed on the identified proteins to examine their role as a potential mediator between social support and CVD and mortality risk.

**Results:** Over a median of 15.8-year follow-up, 889 participants have died, and 627 developed CVD. Of 276 proteins measured, greater social support was associated with lower levels of 13 proteins (EFNA4, SKR3, TNFRSF10A, TNFRSF11A, TRAIL-R2, Gal-9, FGF-23, REN, VSIG2, AMBP, MMP12, ASGR1, PSG1) and higher TN-R levels, after adjusting for baseline socioeconomic confounders. Of these 13 proteins, six proteins (TNFRSF10A, TNFRSF11A, FGF-23, VSIG2, AMBP, and ASGR1) were significantly associated after full adjustments. We also identified 49 protein-CVD and 70 protein-mortality associations after minimal adjustments, including 11 and 14 proteins simultaneously associated with social support. In the mediation analysis, each protein showed significant indirect effects: all the significant proteins together mediated about 20.9% and 26.4 % of the associations for CVD and mortality, respectively. The main enriched biological pathways involved death receptor activity and carbohydrate binding.

**Conclusions:** Social support was related to proteomic signatures and these proteins mediated the association between social support and CVD and mortality risk, independently and cumulatively. These findings deepen our understanding of the intricate connections between relationship quality, proteins, and CVD and mortality development.

## Introduction

Cardiovascular diseases (CVD) are one of the leading causes of morbidity and mortality worldwide ^1^. According to the global disease burden in 2022, there were 621 million prevalent cases of CVD and 24.5 million attributed deaths contributing substantially to the global burden of disease ^1^. Undercovering the modifiable risk factors and the underlying mechanisms is critically important to reducing the risk of morbidity and mortality.

Over the past two decades, substantial advances have been made in understanding how social connections influence morbidity and mortality ^2^. However, much of this work has focused on just two measures of social connections: social isolation and loneliness. While social isolation (a structural component of social connections) refers to the objective presence or absence of contact with others and loneliness (a functional component) refers to subjective assessments as to whether these contacts fulfil individual needs, we understand much less about how other aspects of social connections influence health. One key construct is social support, which is broadly defined as the perception of being cared for and having available assistance when needed ^3^ ^4^, including from family members, close friends, or the community ^5^. Social support has been evidenced to be protective against diverse social outcomes (including isolation and loneliness), psychological outcomes (e.g. depression, and anxiety) and clinical outcomes (including CVD and mortality) ^6–9^. These effects have been demonstrated to be partly achieved through mechanisms including offering psychological resources, helping mitigate stress, and reinforcing health-promoting behaviors ^10^. Theoretically, these results align with hypotheses put forward in socioemotional selectivity theory (SST), which posits that having meaningful social ties becomes more important as people age and is key to experiencing positive wellbeing.

However, we have relatively little understanding about potential underlying biological mechanisms. Further, there are some overlaps between the functional construct of social support and the quality-related construct of relationship strain: if an individual experiences criticism from others or feels they cannot rely on them in times of need, this might not just indicate a lack of social support but also a negative type of interaction that could cause stress and negative emotions ^2^. According to strength and vulnerability integration theory (SAVI), older adults may have reduced physiological flexibility as they age, leading to increased vulnerability to stressors (such as relationship strain) and greater adverse biological effects ^11^. But whether protective effects of social support or potential adverse effects due to relationship strain experienced as part of low social support are more important mechanistically for morbidity and mortality – and biologically how these effects are achieved - remains unclear.

Proteins are the final products of gene expression and serve as the main functional components of biological processes such as signaling pathways, metabolic regulation and immune response ^12^. Meta-analyses of cross-sectional and prospective studies have previously reported associations between social support and various inflammatory proteins such as C reactive protein (CRP), interleukins such as IL-6 and tumour necrosis factor (TNF) as well as fibrinogen ^13^. However, these studies have been focusing on pre-specified individual biomarkers, which limits our broader understanding of the molecular signature of social support. We lack large-scale molecular epidemiology studies exploring the effect of social support on much wider protein panels. This is important if we are to move beyond well-recognised inflammatory biomarkers, look more broadly at potentially relevant biological pathways, and start considering the implications of connected patterns of protein expression rather than single variables. Research into other aspects of social connections such as loneliness and social isolation using molecular epidemiology approaches has been illuminating, identifying specific protein signatures and demonstrating how associated proteins are implicated in clinical conditions such as CVD and mortality ^14,15^.

Consequently, in this current study, we explored several interconnected research questions: (i) What is the plasma proteomic signature of social support and is there evidence of a linear dose-response relationship in levels of social support and protein abundance? (ii) Is this relationship driven more by potential protective effects of social support or adverse effects of relationship strain? (iii) How does the plasma proteomic signature of social support relate to plasma proteomics signatures already identified for other social constructs such as loneliness and isolation and plasma proteomics signatures for CVD and mortality? (iv) Do proteins associated with social support mediate the relationship between social support and CVD and mortality? To answer these questions, we leveraged data from over 3,000 older adults who had proteomics data gathered quantified using the large-scale Olink proteomics platform utilized in the English Longitudinal Study of Ageing (ELSA).

## Methods

This study used data from the ELSA, which is an ongoing longitudinal study of individuals in England aged 50 and older. ELSA is a nationally representative cohort of older men and women living in England ^16^. ELSA collected data from the Health Survey in England (1998, 1999, and 2001) at baseline and had first wave of data in 2002/2003 through in-person interviews and self-reported questionnaire, with biennial follow-up waves. Blood samples were collected starting from the second nurse visit in 2004 (wave 2) and then at four-year intervals thereafter. Ethical approval for ELSA was obtained from the National Research Ethics Service. All participants have provided informed consent.

In the present study, we used wave 4 (2008-2009) as the baseline data in the main analysis because this wave had the proteomic data and the largest number of social support information. Of the 11,050 participants in wave 4 of the ELSA, we excluded those without the assay results of proteins (n= 7,868), those who did not consent the linkage to the health record (n=85) and those aged <50 years (n=36), resulting in a final sample of 3,141 in the analysis of relationship quality and mortality. Then, the participants with CVD at baseline (n= 249) were further excluded, generating the sample of 2,892 for the analysis of association with CVD (Supplemental Figure 1).

### Social support

Questions on social support were administered via self-completion questionnaire and covered four relationship types: (i) spouse/partner; (ii) children; (iii) extended family members; and (iv) friends. Three items examined the participants’ perception of positive aspects of the social support: (i) “How much do they really understand the way you feel about things?”; (ii) “How much can you rely on them if you have a serious problem?”; and (iii) “How much can you open up to them if you need to talk about your worries?”. These items cover empathy, dependability, and confiding, respectively. The other three items examined negative social support: (i) “How much do they criticize you?”; (ii) “How much do they let you down when you are counting on them?”; and (iii) “How much do they get on your nerves?”. These items cover criticism, being let down, and annoyance, respectively. Responses ranged from “not-at-all” (scored 0) to “a lot” (scored 3). Scores from each relationship were calculated separately using the average of the 3 items (1 or 2 items in the case of item missingness).

Both scores of positive and negative social support were calculated by averaging the score from each relationship, with higher scores showing higher positive and negative social support. When calculating the global score, the items’ scores of negative social supports were first reversed and then a global score was calculated by averaging the 8 scores, with higher scores indicating higher social support. However, given some theoretical overlap between social support and relationship strain, we also split the index into positive social support and negative social strain for sensitivity analyses.

### Measurement of health outcomes

The mortality was ascertained by linking the consenting ELSA participants to the National Health Service’s Central Registry, which provides vital status data. CVD events (defined as coronary heart disease (CHD), stroke, and heart failure), were ascertained by linkage to Hospital Episode Statistics data Admitted Patient Care (HES APC) data and HES Outpatient (HES OP) data, by using the International Classification of Diseases, Ninth and Tenth Revision (ICD-10; I20–25, I60–64, I50). For the occurrence of CVD, events were censored at first CVD event of CHD, stroke, or heart failure, whichever came first. The most recent date of mortality was November 2024, and CVD was March 2024. This provided follow up of up to 16 years.

### Proteomic data

The plasma sample to measure proteomic profiles were collected in wave 4 in 2008-09 during nurse visits. The proximity extension assay developed by Olink company was used to measure the proteins. The proteins were detected by binding to two oligonucleotide-linked antibodies, resulting in proximity, and then quantified by next-generation sequencing. Three Olink™ Target 96 panels were selected for the proteomics assays (Cardiovascular II, Neurology I, and Neurology Exploratory), covering a total of 276 unique proteins. These assays incorporate a stringent quality control pipeline has been previously described ^17^.

### Covariates

Directed Acyclic Graph (DAG) was used to identify the potential covariates in the multiple-adjusted models by using the DAGitty online tool. Identified confounders included age, sex, ethnicity, wealth, educational attainment, Body mass index (BMI), and lifestyle behaviours (smoking status, alcohol drinking frequency, and physical activity), assessed through the self-reported questionnaire and respondent interviews. Educational qualifications were categorized into four groups: no educational qualifications; education to GCE/O-levels/national vocational qualification (NVQ) 2 (qualifications at age 16); education to NVQ3/GCE/A-levels (qualifications at age 18); higher qualification/NVQ4/NVQ5/degree). Wealth was recorded using the total net non-pension non-housing wealth (including net value of primary residence, business, and non-housing financial wealth), and participants were categorized into five quantiles. Net non-pension wealth is a robust indicator of socio-economic status in the ELSA population ^18^. Based on the two questions “do you ever smoke” and “are you a current smoker”, smoking status was categorized as never, former and current smoker. Alcohol drinking was dichotomised by the frequency of alcohol consumption into more than 5 days a week or less. Participants were asked about the frequency of vigorous, moderate, and mild physical activity (more than once/week, once/week, 1–3 times/month, or hardly ever). Physical activity was grouped into vigorous (vigorous activity more than once a week), moderate (moderate activity more than once a week), and inactive (mild activity or less frequent activity). BMI was calculated using the participant’s height and weight measured during the nurse visit.

### Statistical analysis

The protein levels were normalized by using the rank-based inverse normal transformation and standardized to mean=0 and SD=1. Missing data on proteins was imputed by using random forests, with 30 imputations and 10 iterations selected. The imputation method was used because of the ability to handle both parametric and non-parametric data sets of complex linear and non-linear problems and to examines biologically important low-abundance peptides ^19^. Results from the imputed datasets were pooled using Rubin’s rule ^20^. Sensitivity analyses were performed by using the complete case analysis.

To assess the associations between social support with each protein, we used linear regressions, by pooling the estimates from all imputed datasets. To account for multiple testing, we applied P value adjusted for false discovery rate (FDR) to indicate statistical significance. We constructed two multiple-adjusted linear regression models: a minimally adjusted model 1 which included sociodemographic variables (age, sex, ethnicity, wealth, education) and model 2 which additionally adjusted for health behaviours (alcohol consumption, smoking, BMI, and physical activity). Cox proportional hazards model was used to identify the significant proteins associated with CVD and mortality, with the two multiple-adjusted models applied. Sensitivity analyses tested whether results were similar when using alternative theoretical definitions of social support, splitting it into positive social support (a functional measure) and relationship strain (a quality measure).

We assessed the mediating role of proteins on the association between social support and CVD and mortality, in the presence of the mediator-outcome confounding variables. DAGs were also used to identify the minimum sufficient adjustment set in the mediation analysis, with identified confounders being age, sex, ethnicity, wealth, and education. The potential mediators (proteins) were selected based on the shared proteins that were both significantly associated with social support and outcome in the models that adjusted for these selected set of confounders, because this study is an exploratory analysis of the underline mechanism of social support and CVD and mortality. All the potential mediators (proteins) with significant effects in one protein simple mediation were included in a multiple mediators mediation model to calculate the combined mediating effect. Two models were estimated: a multivariable linear regression model for proteins conditional on social support as exposure and confounders and a multivariable Cox regression model for CVD or mortality on social support, mediators (proteins) and confounders. The 95% confidence intervals of our estimates were generated using the percentile bootstrapping inference method, with 1000 bootstraps in each procedure and a random seed for reproducibility purposes. Mediation analysis was conducted using the ‘CMAverse’ R package ^21^.

Enrichment analysis was used to assess the biological mechanisms that arts and cultural engagement may modulate and indeed, the potential mechanisms linking social support and CVD and mortality. We used the clusterProfiler package and enrichment analysis in R. This approach adopted hypergeometric tests and FDR correction for enrichment analyses. The analyses were conducted using R, version 4.3.2 (R Foundation for Statistical Computing) statistical packages. Two-tailed p-values <0.05 were considered to indicate significance.

## Results

### Participants characteristics

In our sample of 3,143 participants, the overall average age was 63.8 years□(SD=9.4) at baseline, 54.7% were male, 97.0% were of white ethnicity, and most participants had completed higher education (75.4%) and had never smoked (57.2%). During a median of 15.8-year follow-up, there were 889 deaths and 627 incident CVD cases. Compared with those without CVD or mortality, participants who developed CVD or died during the follow-up were more likely to be older, female, current smokers, frequent alcohol drinker (≥5 days/week), had less wealth and lower education level, and be less physically active (Table 1). The social support score was lower in participants with incident CVD or mortality, compared with those without (Table 1).

**Table 1.** Baseline characteristics of participants by cardiovascular disease and all-cause mortality.

|  | Overall | CVD |  | P value | All-cause mortality |  | P value |
| --- | --- | --- | --- | --- | --- | --- | --- |
|  |  | No | Yes |  | No | Yes |  |
| Number of participants | 3141 | 2265 | 627 |  | 2252 | 889 |  |
| Age (years) | 63.75 (9.36) | 62.03 (8.55) | 67.49 (10.02) | <0.001 | 60.55 (6.64) | 71.86 (10.30) | <0.001 |
| Men, n (%) | 1718 (54.7) | 1312 (57.9) | 301 (48.0) | <0.001 | 1264 (56.1) | 454 (51.1) | 0.012 |
| Education (%) |  |  |  | <0.001 |  |  | <0.001 |
| No educational qualifications | 772 (24.6) | 479 (21.2) | 196 (31.4) |  | 457 (20.3) | 315 (35.6) |  |
| education to GCE/O-levels/ NVQ 2 (qualifications at age 16) | 969 (30.9) | 714 (31.6) | 185 (29.6) |  | 711 (31.6) | 258 (29.1) |  |
| NVQ3/GCE/A-levels (qualifications at age 18) | 286 (9.1) | 208 (9.2) | 52 (8.3) |  | 222 (9.9) | 64 (7.2) |  |
| Higher qualification/NVQ4/NVQ5/degree | 1107 (35.3) | 860 (38.0) | 191 (30.6) |  | 858 (38.2) | 249 (28.1) |  |
| Wealth quintile (%) |  |  |  | <0.001 |  |  | <0.001 |
| Quintile 1 | 607 (19.8) | 393 (17.8) | 140 (22.7) |  | 410 (18.7) | 197 (22.4) |  |
| Quintile 2 | 618 (20.1) | 409 (18.5) | 138 (22.4) |  | 382 (17.4) | 236 (26.9) |  |
| Quintile 3 | 616 (20.1) | 450 (20.4) | 128 (20.7) |  | 435 (19.8) | 181 (20.6) |  |
| Quintile 4 | 615 (20.0) | 466 (21.1) | 118 (19.1) |  | 477 (21.8) | 138 (15.7) |  |
| Quintile 5 | 615 (20.0) | 493 (22.3) | 93 (15.1) |  | 489 (22.3) | 126 (14.4) |  |
| Ethnicity: White (%) | 3047 (97.0) | 2203 (97.3) | 610 (97.3) | 0.999 | 2180 (96.8) | 867 (97.5) | 0.340 |
| Smoking status (%) |  |  |  | 0.018 |  |  | <0.001 |
| Never | 1271 (40.6) | 959 (42.5) | 228 (36.4) |  | 988 (44.0) | 283 (31.8) |  |
| Ever | 1432 (45.7) | 998 (44.2) | 300 (47.8) |  | 991 (44.1) | 441 (49.6) |  |
| Current | 431 (13.8) | 301 (13.3) | 99 (15.8) |  | 266 (11.8) | 165 (18.6) |  |
| Alcohol: ≥5 days/week (%) | 704 (23.1) | 515 (23.5) | 141 (23.2) | 0.912 | 496 (22.8) | 208 (23.9) | 0.549 |
| Physical activity (%) |  |  |  | <0.001 |  |  | <0.001 |
| No/Low | 1026 (32.7) | 647 (28.6) | 259 (41.3) |  | 603 (26.8) | 423 (47.6) |  |
| Moderate | 1388 (44.2) | 1021 (45.1) | 268 (42.7) |  | 1028 (45.6) | 360 (40.5) |  |
| High | 727 (23.1) | 597 (26.4) | 100 (15.9) |  | 621 (27.6) | 106 (11.9) |  |
| BMI (mean (SD)) | 27.46 (6.86) | 27.15 (6.28) | 28.30 (8.12) | <0.001 | 27.74 (6.05) | 26.74 (8.55) | <0.001 |
| Social support (mean (SD)) | 3.09 (0.39) | 3.11 (0.39) | 3.04 (0.39) | <0.001 | 3.11 (0.39) | 3.02 (0.39) | <0.001 |
| Positive social support (mean (SD)) | 2.71 (0.81) | 2.77 (0.80) | 2.60 (0.82) | <0.001 | 2.80 (0.78) | 2.50 (0.85) | <0.001 |
| Relationship strain (mean (SD)) | 1.54 (0.43) | 1.55 (0.43) | 1.51 (0.43) | 0.048 | 1.57 (0.42) | 1.45 (0.43) | <0.001 |

Abbreviations: BMI, body mass index; GCE, general certificate of education; NVQ, national vocational qualification; SD, Standard deviation;

### Social support and proteins

We conducted linear regression involving 276 plasma proteins, using social support as the exposure and proteins as the outcome. After adjusting for the baseline socioeconomic confounders, greater social support was significantly associated with lower levels of 13 proteins (EFNA4, SKR3, TNFRSF10A, TNFRSF11A, TRAIL-R2, Gal-9, FGF-23, REN, VSIG2, AMBP, MMP12, ASGR1, PSG1) and higher levels of one protein (TN-R) when considering the FDR-based multiple testing correction (Figure 1a; Figure 2; Supplemental Table 1). VSIG2 demonstrated the strongest association with social support (coefficient (β) [standard error (SE)]: -0.224 [0.044]; FDR adjusted P value (denoted as P_FDR_) = 0.005), followed by FGF_23 (β [SE]: -0.203 [0.046]; P_FDR_ = 0.025), TNFRSF11A (β [SE]: -0.185 [0.044]; P_FDR_ = 0.018), TRAIL_R2 (β [SE]: -0.183 [0.044]; P_FDR_ =0.002) and TRAIL_R2 (β [SE]: -0.174 [0.044]; P_FDR_ =0.005).

**Figure 1.**
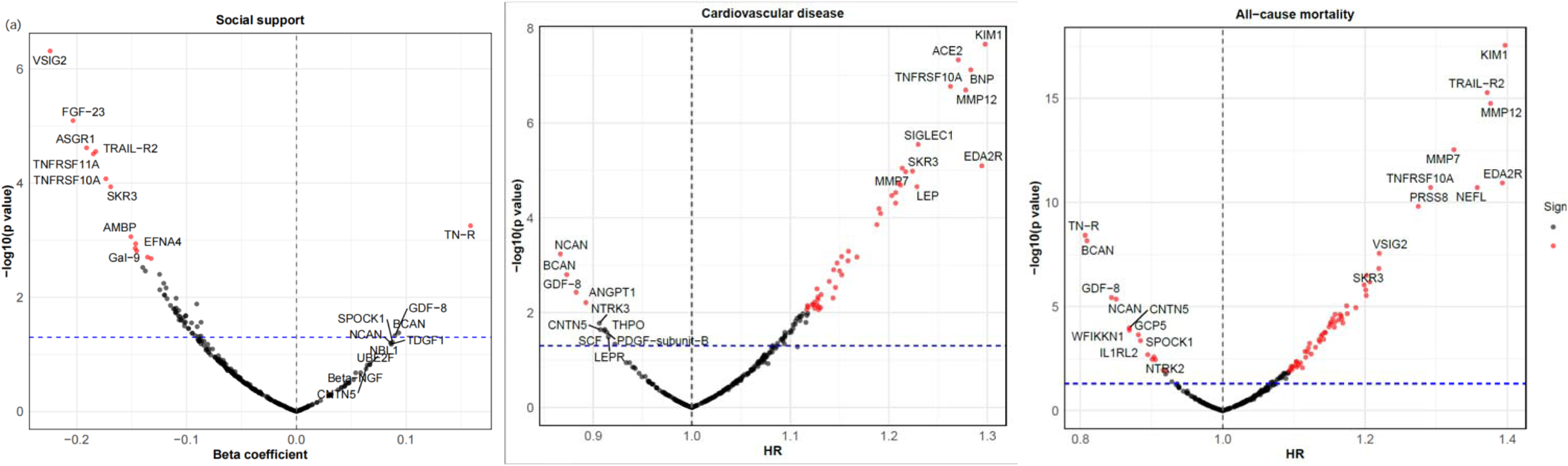
Volcano plots showing proteins associated with social support (a), all-cause mortality (b), CVD (c), highlighting strongly associated proteins. The x axis represents beta coefficient in the linear regression (social support and proteins) or hazard ratio in the cox hazard regression model (proteins and CVD and all-cause mortality), and the y axis represents −log10(P values). The blue dashed lines indicate P value <0.05. The red dots indicate the proteins having the FDR adjusted P value <[0.05 and the grey dots indicate the proteins with FDR adjusted P value larger than 0.05. The correlations were all adjusted for age, sex, education, wealth, and ethnicity.

**Figure 2.**
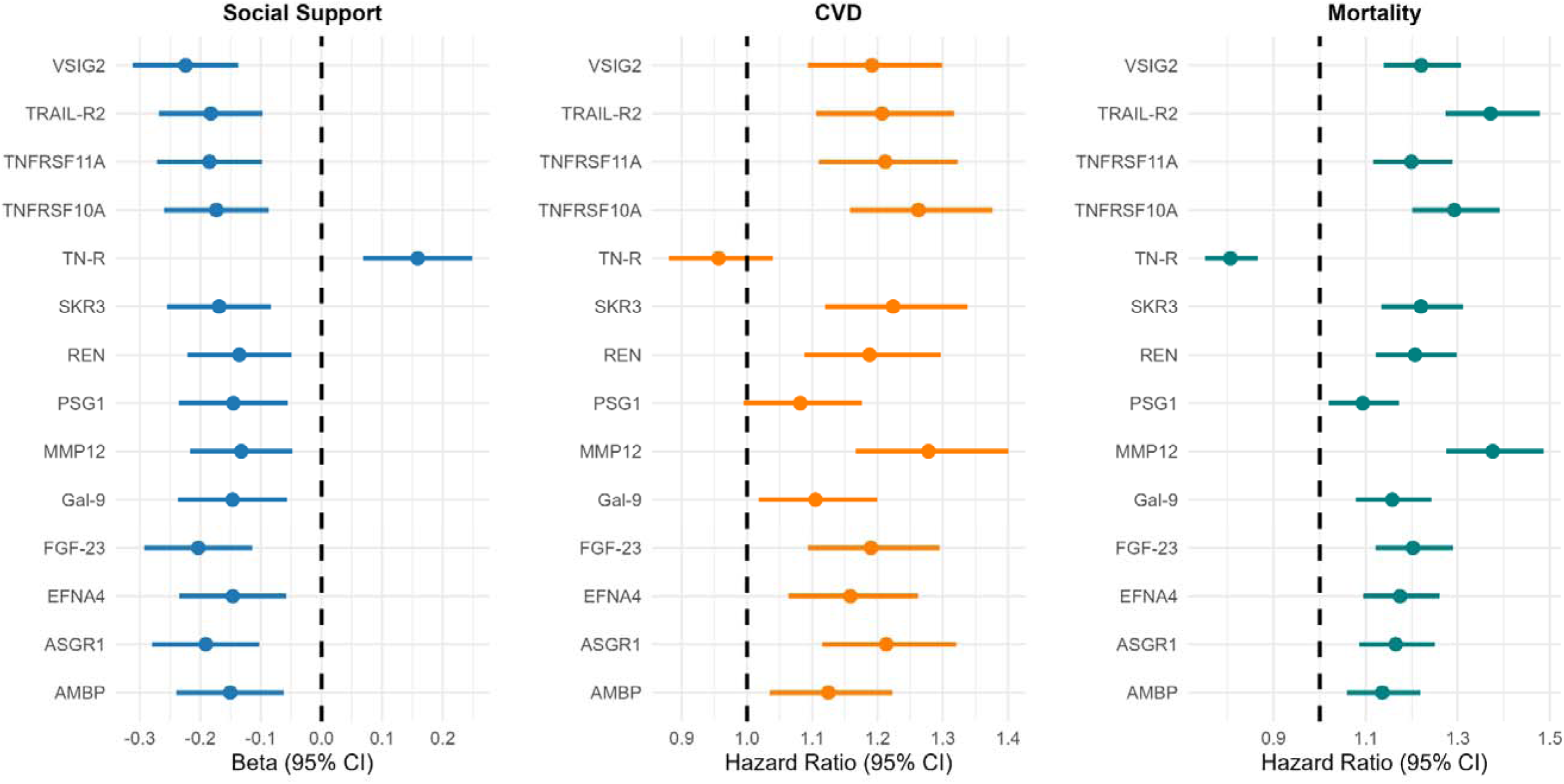
Significant proteins associated with social support and the association with cardiovascular disease and all-cause mortality. The figure only showed the proteins that were significant in the association between social support and proteins with FDR adjusted P value <0.05 and the association of these proteins and CVD and all-cause mortality. Model adjusted for age, sex, wealth, education, and ethnicity.

After further adjusting for smoking status, alcohol use, physical activity, and BMI, six proteins remained significant after full adjustments: TNFRSF10A (β [SE]: -0.156 [0.044]; P_FDR_ =0.020), TNFRSF11A (β [SE]:-0.194 [0.044]; P_FDR_ = 0.003), FGF-23 (β [SE]:-0.182 [0.045]; P_FDR_ = 0.004), VSIG2 (β [SE]:-0.180 [0.044]; P_FDR_ = 0.004), AMBP (β [SE]:-0.150 [0.045]; P_FDR_ = 0.040), and ASGR1 (β [SE]:-0.187 [0.044]; P_FDR_ = 0.003) (Supplemental Figure 2a; Supplemental Table 1). Restricted cubic splines revealed no evidence of the non-linear relationship for 12 proteins and social support (Figure 3).

**Figure 3.**
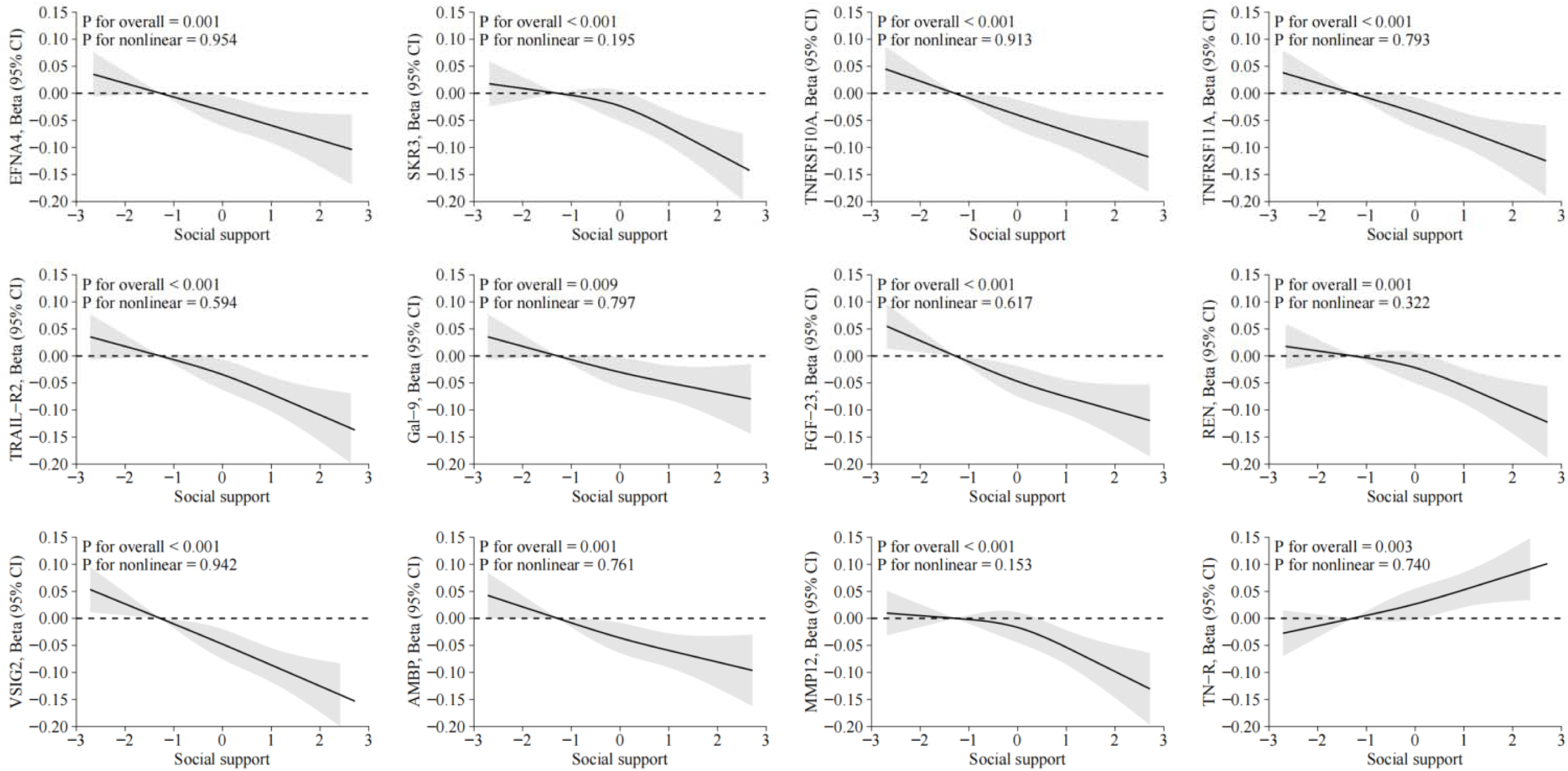
Dose-response relation between social support and protein. The analyses were performed in model that adjusted for age, sex, wealth, education, and ethnicity. The solid lines show beta coefficient and the shaded areas show 95% confidential intervals (CIs).

When splitting the analyses into potential alternative classifications of positive social support and relationship strain, positive social support was significantly associated with lower levels of 13 proteins (EFNA4, SKR3, TNFRSF10A, TNFRSF11A, TRAIL-R2, FGF-23, REN, VSIG2, AMBP, MMP12, FGFR2, NEFL, NXPH1, PSG1) and social strain was associated with lower levels of one protein (LRPR), after adjusting for the minimal covariates (Supplemental Figure 3; Supplemental Table 2). When further adjusting for lifestyle and BMI, 9 proteins remained significant, but 3 proteins became non-significant, and 5 proteins became significant (Supplemental Figure 3; Supplemental Table 2).

### Proteins and CVD and all-cause mortality

Of the 276 proteins explored, Cox Proportional Hazard regression models looking at outcomes over the following 16 years showed 45 proteins were significantly associated with increased risk of CVD and 4 proteins were associated with decreased risk in model 1, which adjusted for age, sex, ethnicity, wealth, and education (Figure 2b; Supplemental Table 5). After further adjusting for smoking status, alcohol use, physical activity, and BMI (model 2), 18 proteins were significantly associated with risk of incident CVD (UNC5C, MSR1, SIGLEC1, GFR_alpha_1, SKR3. EDA2R, TNFRSF10A, TNFRSF11A, FGF-23, KIM1,

VSIG2, MMP7, MMP12, ACE2, CTSL1, IKZF2, RPS6KB1) (Supplemental Figure 2b; Supplemental Table 3). The strongest association was BNP (HR: 1.286; 95% CI 1.175,1.407; P_FDR_ <0.001), followed by EDA2R, KIM1, MMP12, ACE2, TNFRSF10A, SIGLEC1, MSR1, SKR3, and MMP7 (Supplemental Table 3).

For all-cause mortality over the following 16 years, Cox Proportional Hazard regression models showed positive association with 57 proteins and negative association with 13 proteins, after adjustment of age, sex, ethnicity, wealth, and education (Figure 2c). After further adjusting for smoking status, alcohol use, physical activity, and BMI, 55 proteins remained significantly associated with all-cause mortality risk, with EDA2R showing strongest estimate (HR: 1.362; 95% CI 1.239,1.496; P_FDR_ <0.001), followed by NEFL, KIM1, MMP12, TRAIL_R2, MMP7, TNFRSF10A, PRSS8, TNFRSF11A, and REN (Supplemental Figure 2c; Supplemental Table 4).

### Mediation analysis

Social support was significantly associated with a lower risk of CVD over the following 16 years (HR 0.747; 95% CI 0.609,0.918) and mortality (0.718; 0.602,0.857) in the full adjustment model (Supplemental Table 5). Among these significant proteins associated with CVD, 12 proteins were also related to social support in the minimally adjusted models (Figure 2; Supplemental Table 6; Supplemental Figure 4) and were further explored whether social support decreased risk of CVD through these proteins. The mediation analysis showed that VSIG2 contributed to the largest proportion of mediation analysis (10.2%), followed by TNFRSF10A (10.0%), ASGR1 (9.1%), TNFRSF11A (8.8%), and FGF-23 (8.7%). Collectively, the 12 proteins significantly and partially mediated 20.9% (3.3% to 38.5%) of the social support-CVD association (Table 2).

**Table 2.**
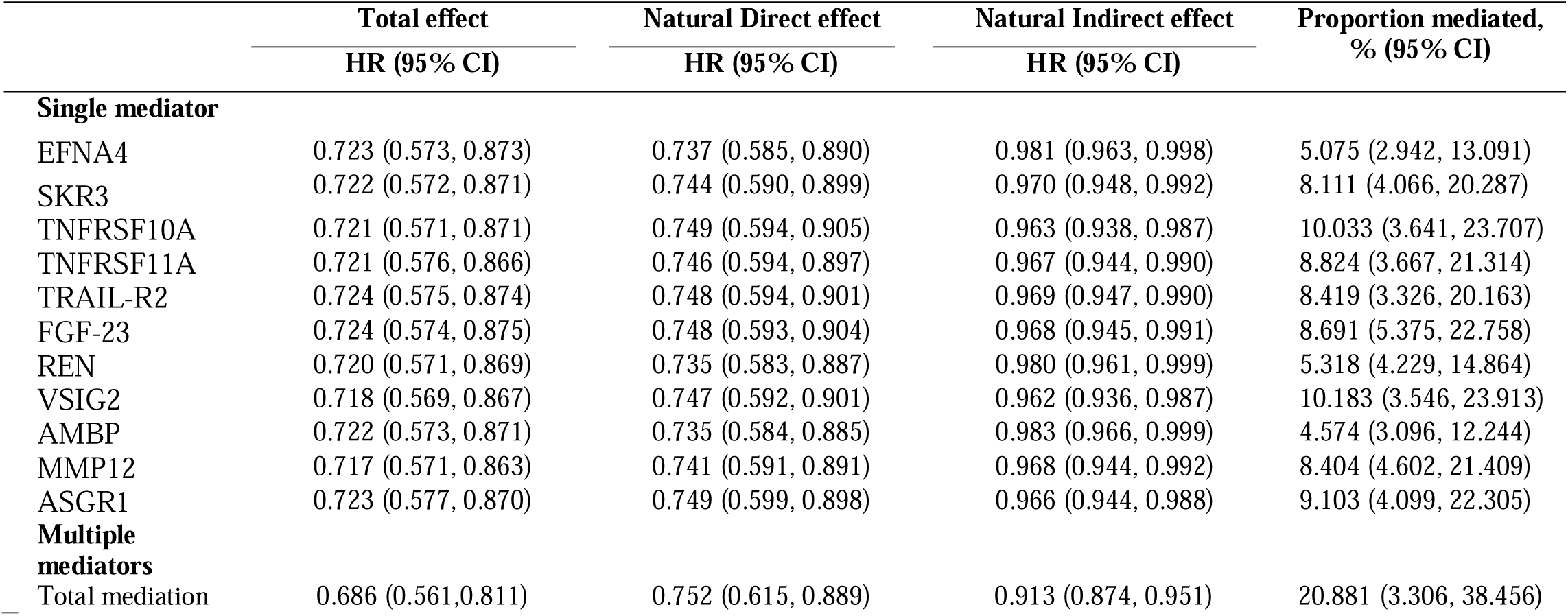
Exploration of potential protein mediators in the association of social support and cardiovascular disease.

|  | Total effect | Natural Direct effect | Natural Indirect effect | Proportion mediated, |
| --- | --- | --- | --- | --- |
|  | HR (95% CI) | HR (95% CI) | HR (95% CI) | % (95% CI) |
| **Single mediator** |  |  |  |  |
| EFNA4 | 0.723 (0.573, 0.873) | 0.737 (0.585, 0.890) | 0.981 (0.963, 0.998) | 5.075 (2.942, 13.091) |
| SKR3 | 0.722 (0.572, 0.871) | 0.744 (0.590, 0.899) | 0.970 (0.948, 0.992) | 8.111 (4.066, 20.287) |
| TNFRSF10A | 0.721 (0.571, 0.871) | 0.749 (0.594, 0.905) | 0.963 (0.938, 0.987) | 10.033 (3.641, 23.707) |
| TNFRSF11A | 0.721 (0.576, 0.866) | 0.746 (0.594, 0.897) | 0.967 (0.944, 0.990) | 8.824 (3.667, 21.314) |
| TRAIL-R2 | 0.724 (0.575, 0.874) | 0.748 (0.594, 0.901) | 0.969 (0.947, 0.990) | 8.419 (3.326, 20.163) |
| FGF-23 | 0.724 (0.574, 0.875) | 0.748 (0.593, 0.904) | 0.968 (0.945, 0.991) | 8.691 (5.375, 22.758) |
| REN | 0.720 (0.571, 0.869) | 0.735 (0.583, 0.887) | 0.980 (0.961, 0.999) | 5.318 (4.229, 14.864) |
| VSIG2 | 0.718 (0.569, 0.867) | 0.747 (0.592, 0.901) | 0.962 (0.936, 0.987) | 10.183 (3.546, 23.913) |
| AMBP | 0.722 (0.573, 0.871) | 0.735 (0.584, 0.885) | 0.983 (0.966, 0.999) | 4.574 (3.096, 12.244) |
| MMP12 | 0.717 (0.571, 0.863) | 0.741 (0.591, 0.891) | 0.968 (0.944, 0.992) | 8.404 (4.602, 21.409) |
| ASGR1 | 0.723 (0.577, 0.870) | 0.749 (0.599, 0.898) | 0.966 (0.944, 0.988) | 9.103 (4.099, 22.305) |
| **Multiple mediators** |  |  |  |  |
| Total mediation | 0.686 (0.561, 0.811) | 0.752 (0.615, 0.889) | 0.913 (0.874, 0.951) | 20.881 (3.306, 38.456) |

Among the significant proteins of all-cause mortality, 14 proteins were found to be associated with social support in the minimally adjusted models (Figure 2; Supplemental Table 6; Supplemental Figure 4) and were further explored whether social support decreased risk of all-cause mortality through these proteins. The proportion mediated by TRAIL-R2 was largest in the single mediation analysis (10.6%), followed by TNFRSF10A (8.1%), VSIG2 (7.9%), MMP12 (7.7%), and FGF-23 (6.7%); all the 14 proteins significantly and partially mediated a total of 26.4% (7.7% to 45.1%) of the social support-all-cause mortality association (Table 3).

**Table 3.** Exploration of potential protein mediators in the association of social support and all-cause mortality.

|  | Total effect | Natural Direct effect | Natural Indirect effect | Proportion mediated, |
| --- | --- | --- | --- | --- |
|  | HR (95% CI) | HR (95% CI) | HR (95% CI) | % (95% CI) |
| **Single mediator** |  |  |  |  |
| EFNA4 | 0.654 (0.535, 0.773) | 0.668 (0.547, 0.790) | 0.978 (0.961, 0.995) | 4.245 (0.222, 8.711) |
| SKR3 | 0.652 (0.534, 0.770) | 0.673 (0.552, 0.795) | 0.969 (0.948, 0.989) | 6.048 (0.435, 11.661) |
| TNFRSF10A | 0.649 (0.532, 0.767) | 0.678 (0.555, 0.800) | 0.958 (0.934, 0.981) | 8.123 (1.481, 14.766) |
| TNFRSF11A | 0.649 (0.531, 0.767) | 0.670 (0.548, 0.792) | 0.969 (0.949, 0.988) | 5.977 (0.618, 11.336) |
| TRAIL-R2 | 0.648 (0.531, 0.765) | 0.685 (0.562, 0.808) | 0.946 (0.918, 0.974) | 10.576 (2.062, 19.09) |
| FGF-23 | 0.651 (0.533, 0.768) | 0.674 (0.552, 0.796) | 0.965 (0.944, 0.986) | 6.702 (0.752, 12.652) |
| REN | 0.655 (0.535, 0.775) | 0.671 (0.549, 0.793) | 0.976 (0.958, 0.994) | 4.629 (0.016, 9.273) |
| VSIG2 | 0.648 (0.530, 0.767) | 0.676 (0.553, 0.799) | 0.959 (0.938, 0.98) | 7.936 (1.524, 14.349) |
| AMBP | 0.651 (0.535, 0.767) | 0.663 (0.544, 0.781) | 0.982 (0.967, 0.997) | 3.396 (0.303, 7.094) |
| MMP12 | 0.641 (0.524, 0.758) | 0.668 (0.548, 0.788) | 0.959 (0.932, 0.987) | 7.551 (0.904, 14.198) |
| ASGR1 | 0.652 (0.531, 0.772) | 0.669 (0.546, 0.793) | 0.973 (0.955, 0.991) | 5.127 (0.049, 10.302) |
| Gal-9 | 0.657 (0.538, 0.775) | 0.670 (0.549, 0.791) | 0.980 (0.964, 0.996) | 3.851 (0.386, 8.087) |
| PSG1 | 0.656 (0.541, 0.771) | 0.663 (0.547, 0.780) | 0.989 (0.976, 1.001) | 2.186 (0.879, 5.252) |
| TN-R | 0.663 (0.543, 0.782) | 0.684 (0.561, 0.808) | 0.968 (0.947, 0.99) | 6.400 (0.155, 12.645) |
| **Multiple mediators** |  |  |  |  |
| **Total mediation** | 0.653 (0.531, 0.776) | 0.745 (0.607, 0.883) | 0.877 (0.834, 0.920) | 26.406 (7.673, 45.139) |

### Results from complete case analysis

Sensitivity analyses using complete case analysis of proteins were performed as the sensitivity analysis to test the robustness of the association of social support, CVD and mortality with plasma protein levels and the potential mediation role of proteins. The analysis yielded results consistent with the main analysis by multiple imputations using random forest. After adjusting for age, sex, wealth, education, and ethnicity, 13 proteins were associated with social support except for Gal-9 showing non-significant association (Supplemental Table 7). Of 13 proteins, 11 and 12 proteins were also associated with increased risk of CVD and mortality, respectively, and 1 protein (TN-R) showed inverse association with mortality. The mediation analysis results were qualitatively similar to those from the imputed data, showing that these proteins significantly and partially mediated 25.6% (11.0% to 83.2%) and 29.4% (17.2% to 57.2%) of the social support-CVD and -mortality association, respectively (Supplemental Tables 8 and 9).

### Pathway enrichment

Enrichment analysis for Gene Ontology (GO) Biological Process, Molecular Function, Kyoto Encyclopedia of Genes and Genomes (KEGG), Reactome, WikiPathways were performed on the proteins significantly linked to social support. The top functional enrichments in social support-associated proteins are presented in Figure 4 and Supplemental Table 10. GO-Molecular Function enrichment analyses showed proteins were robustly enriched into those pathways mainly related to cell survival/apoptosis (death receptor activity, insulin-like growth factor receptor binding, ephrin receptor activity), immune function (IgA binding, immunoglobulin binding, fucose binding, carbohydrate binding, D-mannose binding, tumor necrosis factor receptor activity), atherosclerosis/vascular and metabolism (Ephrin receptor activity, calcium channel inhibition, IGF signaling, calcium oxalate binding, asialoglycoprotein receptor, insulin-like growth factor receptor binding), inflammation (TRAIL binding, TNF receptor activity, aspartic-type peptidase activity, endopeptidase activity) and other pathways (Transmembrane signaling receptor activity, molecular transducer activity).

**Figure 4.**
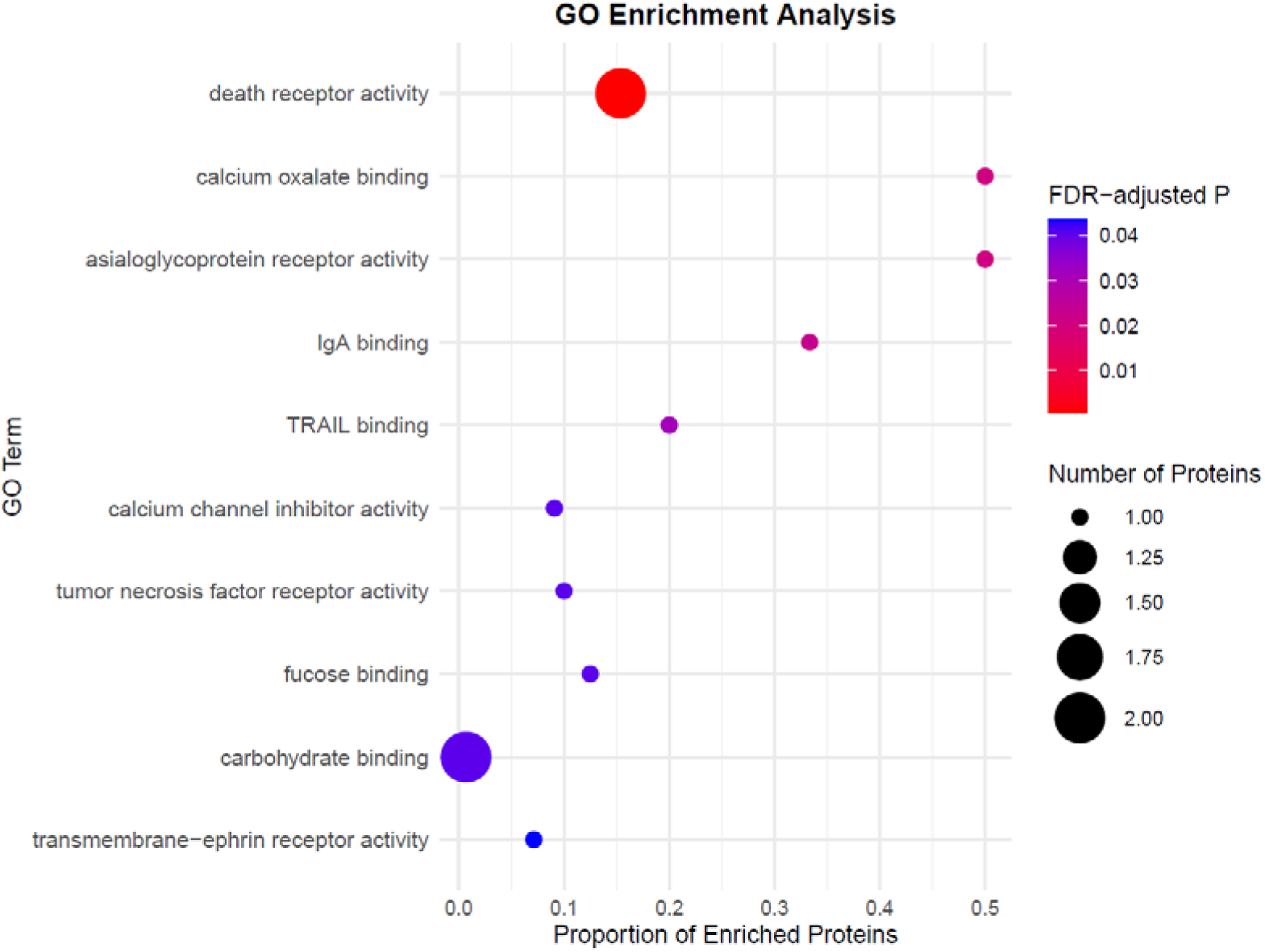
GO enrichment analyses of the 14 proteins associated with social support. Rows show the GO terms, the dot sizes show the number of enriched proteins, the colors indicate the FDR-adjusted *P* value and the *x* axis shows the proportion of enriched proteins relative to all proteins associated with the GO term.

## Discussion

In this study, we unravel the associations between social support and a vast array of circulating proteins and the mediation role of proteins in the association between social support and CVD and all-cause mortality. The proteome analyses identified 13 positive and 1 inverse protein-social support associations, presenting a linear dose-response relationship. These proteins provide clear extensions beyond any previously researched in relation to social support, which is important in identifying novel biological pathways ^12^. All these 14 proteins significantly and partially mediated the prospective association between social support and all-cause mortality while controlling for covariates (accounting for 26.4% of the effects); 12 of them were also related to incident CVD and partially mediated the prospective association with social support (accounting for 20.9% of the effects).

The biological mechanisms underlying the association between social relationships and health remain a large knowledge gap ^22^. Profiling protein in relation to social support could provide direct insights into disease pathophysiology. Previous studies have found the significant association between social isolation and loneliness and protein signatures ^14^ ^15^. Zhang et al. (2025) ^14^ used 42,062 participants across 2,920 plasma proteins in the UK Biobank to explore the plasma proteomic signatures of social isolation and loneliness and they identified 776 proteins significantly associated with social isolation and 519 proteins associated with loneliness. The previous study in our team by Gong et al. (2025) ^15^ also identified 11 proteins associated with social isolation but no proteins significantly associated with loneliness, using ELSA. Our study extends the research findings to explore the protein signatures of social support. To some extent, there were some overlaps and distinction between the proteins associated with social isolation and those associated with social support identified in the present study. As shown in the supplemental figure 5, compared with studies by Gong et al. (2025) using the same dataset (ELSA) ^15^, 7 common proteins are identified for both social support and social isolation (SKR3, TNFRSF10A, TNFRSF11A, TRAIL-R2, REN, VSIG2, MMP12) and 7 proteins (EFNA4, Gal-9, FGF-23, AMBP, ASGR1, PSG1, TN-R) were distinctly associated with social support. The UK Biobank study showed some shared proteins, including EFNA4, FGF23, AMBP, TNFRSF10A, TNFRSF11A, and ASGR1 ^14^. Discrepancy may stem from the different measurements of social isolation and population characteristics or simply due to FDR adjustment as UK Biobank tested much wider panels of proteins. Further, our findings corroborate some findings in previous epidemiological analyses focusing on specific biomarkers, such as recurrent finding of proteins that are part of the TNF superfamily ^12^.

We also considered different theoretical conceptualisations of social support vs relationship strain, finding that the associations were almost entirely driven by the protective effects of social support rather than the adverse effects of social strain. This is particularly in line with socioemotional selectivity theory, suggesting that it is the positive and meaningful aspects of social connections such as perceived positive social support contribute beneficially towards health, rather than negative aspects such as relationship strain adversely affecting health ^23^. This echoes other biological research on physiological age acceleration, which similarly reported that social support has stronger associations with physiological age than social strain ^24^. It also aligns with research looking at stress-related neuroendocrine pathways, which similarly found protective effects from positive social support than adverse effects from relationship strain ^25^. Notably, both studies also used ELSA data and the same social questions, providing clear methodological alignment.

Cumulative studies have shown social support plays a critical role in the health ^9^. Consistent with previous studies ^26,27^, the present study showed the strong association between social support and CVD and all-cause mortality, where each 1-point increase in the social support score was associated with a more than 2.5-fold increase in the hazard of CVD and mortality. We identified novel proteins that may be influenced by social support and found all proteins with a significant relationship with social support were prospectively associated with the incidence of CVD and mortality and partially mediated the association between social support and these outcomes. Specifically, social support was negatively associated with the levels of VSIG2, TRAIL-R2, TNFRSF10A, TNFRSF11A, and Gal-9, suggesting that inflammation and apoptosis may be important biological pathways. VSIG2 (V-set and immunoglobulin domain-containing protein 2), a protein belonging to the immunoglobulin superfamily and playing a role in immune regulation ^28^, was observed the strongest association. TRAILR2, known as death receptor 5, is a receptor belonging to the TNF superfamily that preferentially regulates apoptosis ^29^. TRAIL-R2 is also found on endothelial cells and vascular smooth muscle cells and the interaction with TRAIL can lead to activation of inflammation signals and induce vascular inflammation ^30^. High levels of TRAIL-R2 have been associated with adverse cardiovascular events and mortality in the population ^31^. TNFRSF10A, known as TRAIL-R1 or death receptor 4, is a receptor for TRAIL (TNF-related apoptosis-inducing ligand) and primarily mediates apoptosis ^32^. TNFRSF11A, known as receptor activator of nuclear factor kappa-B (RANK), is receptor inducing the activation of NF-kappa B and MAPK8/JNK. It plays a key role in immune cell development, and inflammatory responses and has been shown in various diseases such as cancer, metabolic disease and CVD ^33–35^. Gal-9, as a member of the galectin family, is a receptor on the various immune cell and regulates the immune responses ^36^, which is involved in various inflammatory diseases such as rheumatoid arthritis ^37^ and also shown to be a target for cancer immunotherapy and a biomarker of cancer ^38^ and CVD ^39^. PSG1 is the family of pregnancy specific glycoproteins ^40^ and activates the latent form of total TGF-β1and increased its secretion ^41^, which plays a crucial role in inflammation. What is notable across all these findings is that high social support does not necessarily activate protective biological pathways, but rather reduces levels of potentially damaging biological responses.

We also found the significant mediation role of FGF-23, EFNA4, REN, ASGR1 and MMP12 in the relationship between social support and CVD and mortality. Previous studies have found increased levels of FGF-23 ^42–45^, ASGR1 ^45,46^, REN ^45–48^, MMP12 ^45,46,49,50^, EFNA4 ^46^, AMBP ^51^ were associated with CVD or mortality, which also support the mediation findings in the present study. The findings may indicate atherosclerosis/vascular and metabolism-related pathways as an important pathway of social support-health relation. FGF-23 is an endocrine hormone that has a key role in phosphate and calcium metabolism ^52^ and decreases circulating concentrations of 1,25-dihydroxyvitamin D ^53^. REN is a highly specific endopeptidase and involved in the activation of renin-angiotensin aldosterone system (RAAS) and contribute to high blood pressure ^54^, vascular inflammation and injury ^55^. EFNA4, known as Ephrin-A4, is involved in angiogenesis, axon guidance, and vascular development and emerging evidence has shown the pivotal contribution to cardiovascular physiology and pathology, except for neurogenesis and cancer ^56,57^. AMBP, also known as the alpha-1-microglobulin/bikunin precursor, was also found to be strongly and positively associated with the albumin excretion rate ^58^ and related to cancer and cancer survival ^59,60^.

To our knowledge, this is the first study to explore the relation between social support and plasma protein levels and explore whether these proteins play a pivotal role in mediating the connection between social support and CVD and mortality. Our analysis identified the proteins associated with social support by undertaking a proteome-wide data-driven approach in a nationally representative cohort of older adults and confirmed associations of social support□related proteins with CVD and mortality. Meanwhile, this study has the strength of performing a series of mediation analysis integrated with the Cox hazard regression model, allowing for the decomposition of total effects into direct and indirect effects in the context of time-to-event outcomes. Considering the limited evidence, further research is required to replicate the findings here in other datasets and populations. Despite the strengths, several limitations should be noted when interpreting the results. First, only a small subset of the roughly 20,000 proteins catalogued by the Human Proteome Project were covered by our proteomic assay, which primarily target proteins involved in neurological and cardiovascular processes in only three panels (Cardiovascular II, Neurology I, and Neurology Exploratory). Future study is warranted to cover a wide range of proteomic data to explore the other under-investigated protein biomarkers of social support. Second, the proteins were measured only at one wave, which hindered the ability to evaluate changes in proteins and health. Third, the participants enrolled in the ELSA tend to represent a more health-conscious population, leading to the potential selection bias, although it is notable that rates of CVD and mortality reported are aligned with population estimates. The participants focused on the older people aged above 50 years and were mainly of white ethnicity, so it is worth exploring whether results can be generalised to other study populations. Fourth, our study used cross-sectional data for social support and proteins, although it had longitudinal data for CVD and mortality outcomes. So, the temporality between proteins and social support cannot be fully established. Future extensions of this work are encouraged that could draw on temporal or genetic approaches to improve causal inference. Finally, although multiple confounders have been adjusted for, residual confounding cannot be excluded, which limits the causal inference.

## Conclusions

Overall, social support was related to proteomic signatures and showed a linear dose-response relationship. The effect of social support on CVD and mortality appears to be mediated by these proteins (approximately 25%). This study suggests novel biological pathways beyond those pathways previously most studied experimentally, including those involved in cell survival/apoptosis, immune function, atherosclerosis and metabolism, transmembrane signalling receptor activity, and molecular transducer activity. These pathways and the specific proteins identified may function as causal mechanisms underpinning the relationship between social support and morbidity and mortality, with support for theoretical propositions that positive aspects of social connections may be protective for health in older age. Our results show some synergy with previous research focused on other aspects of social connections but also highlight novel proteins involved, reinforcing the importance of research exploring diverse aspects of social connections in relation to biological processes and health outcomes in order to gain appropriate nuanced insight into which aspects of social connections may be most appropriate for targeting in psychosocial interventions.

## Acknowledgements

This study was funded by the National Institute on Aging (grant number R01AG17644) and the National Institute for Health and Care Research (198/1074–02).

## Author contributions

P.Q and D.F. contributed to the conception or design of the work, results interpretation and manuscript writing. P.Q and J.G. contributed to data cleansing and analysis; J.G. and A.S. contributed to the acquisition and interpretation of data, and the manuscript review and revision. All authors approved the final manuscript.

## Competing interests

The authors declare no competing interests.

## Data availability

ELSA data are available through registration with the UK data service (https://beta.ukdataservice.ac.uk/datacatalogue/series/series?id=200011). The proteomics data in ELSA will be deposited on the UK Data Service upon publication.

## Notes

### Competing Interest Statement

The authors have declared no competing interest.

### Author Declarations

Ethics committee of the National Research Ethics Service gave ethical approval for this work.

