## Supplemental Tables and Figures for "Plasma proteomic signatures of social support and their association with cardiovascular disease and mortality"

### Supplemental material

**Supplemental Figure 1** Participants selection

**Supplemental Figure 2** Volcano plots showing proteins associated with social support (a), CVD (b), all-cause mortality (c) in model 2

**Supplemental Figure 3** Volcano plots showing the correlation between protein and positive social support in model 1 (a) and in model 2 (b), and relationship strain in model 1 (c) and in model 2 (d).

**Supplemental Figure 4** Proteins are shared and distinct for the associations between protein signatures with social support (linear regression analysis), and all-cause mortality (Cox regression) and CVD (Cox regression).

**Supplemental Figure 5** Venn diagram visualizing the overlap of measured protein levels associated with social support after correction and the proteins associated with social isolation shown in the study by Gong, et al. (2025) and that by Shen, et al. (2025).

**Supplemental Table 1** Associations between social support and proteins

**Supplemental Table 2** Associations between positive social support, relationship strain and proteins

**Supplemental Table 3** Associations between proteins and cardiovascular disease

**Supplemental Table 4** Associations between proteins and all-cause mortality

**Supplemental Table 5** Association of social support and cardiovascular disease and all-cause mortality

**Supplemental Table 6** Association of protein and social support, cardiovascular disease and all-cause mortality

**Supplemental Table 7** Sensitivity analyses using complete case analysis for the association of protein and social support, cardiovascular disease and all-cause mortality

**Supplemental Table 8** Sensitivity analyses using complete case analysis for the potential protein mediators in the association of social support and cardiovascular disease

**Supplemental Table 9** Sensitivity analyses using complete case analysis for exploration of potential protein mediators in the association of social support and all-cause mortality

**Supplemental Table 10** Significant functional enrichments in social support-associated proteins

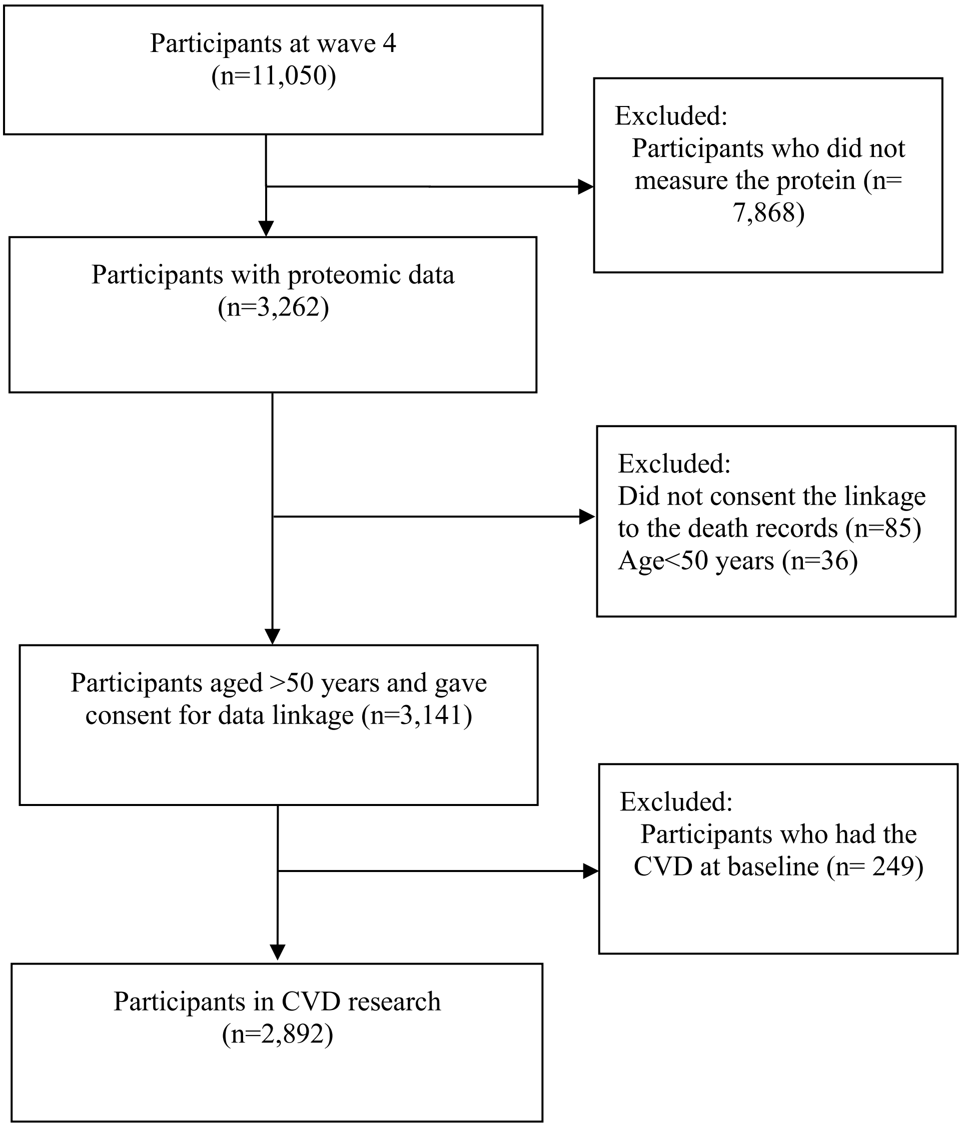
Participants at wave 4
(n=11,050)
Excluded:
Participants who did not measure the protein (n= 7,868)
Participants with proteomic data
(n=3,262)
Excluded:
Did not consent the linkage to the death records (n=85)
Age<50 years (n=36)
Participants aged >50 years and gave consent for data linkage (n=3,141)
Excluded:
Participants who had the CVD at baseline (n= 249)
Participants in CVD research
(n=2,892)

**Supplemental Figure 1 Participants selection**

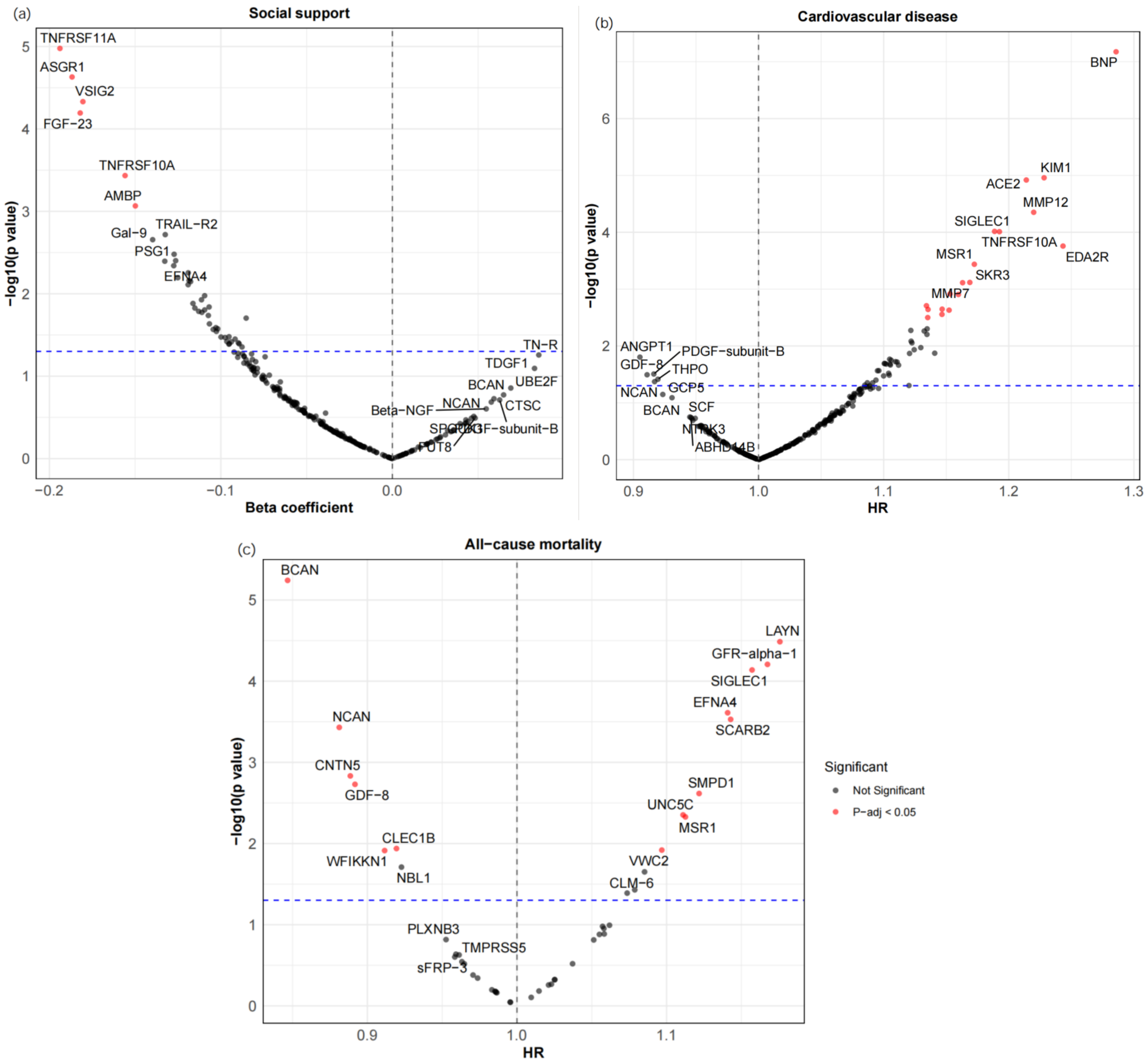


**Supplemental Figure 2 Volcano plots showing proteins associated with social support (a), CVD (b), all-cause mortality (c) in model 2**

The x axis represents beta coefficient in the linear regression (social support and proteins) or hazard ratio in the cox hazard regression model (proteins and CVD and all-cause mortality), and the y axis represents −log10(P values). The blue dashed lines indicate P value <0.05. The red dots indicate the proteins having the FDR adjusted P value < 0.05 and the grey dots indicate the proteins with FDR adjusted P value larger than 0.05. The correlations were all adjusted for age, sex, wealth, education, ethnicity, smoking status, alcohol use, physical activity, and BMI.

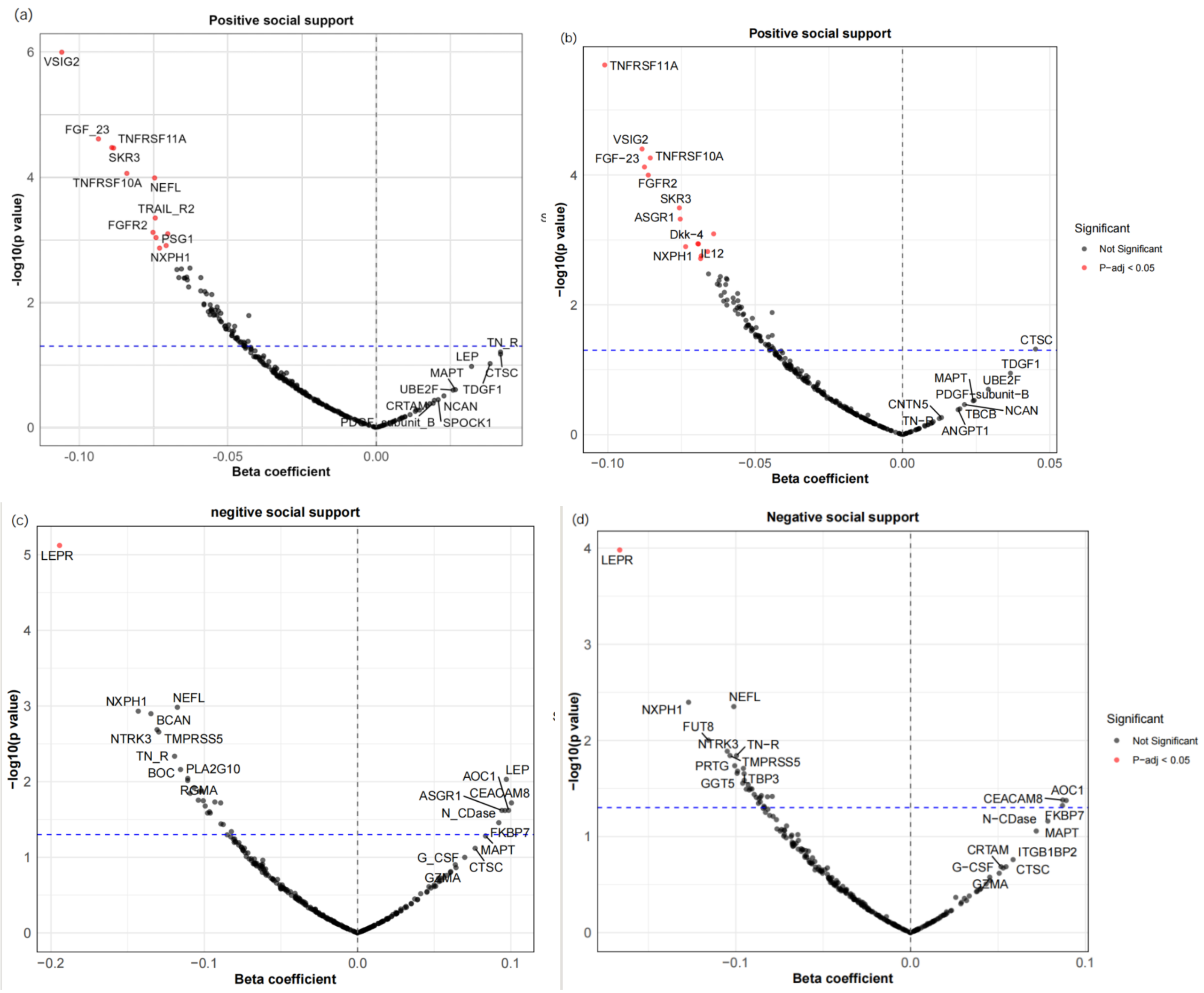


**Supplemental Figure 3 Volcano plots showing the correlation between protein and positive social support in model 1 (a) and in model 2 (b), and relationship strain in model 1 (c) and in model 2 (d).**

The x axis represents beta coefficient, and the y axis represents −log10(P values). The blue dashed lines indicate P value <0.05. The proteins that marked red indicate the proteins having the FDR adjusted P value < 0.05.

Model 1: adjusted for age, sex, wealth, education, and ethnicity.

Model 2: additionally adjusted for smoking status, alcohol use, physical activity, and BMI.

#### CVD

**Social support**
**11 proteins**
**EFNA4, SKR3, TNFRSF10A, TNFRSF11A, TRAIL-R2, FGF-23, REN, VSIG2, AMBP, MMP12, ASGR1**

**38 proteins**
**UNC5C, CLM-6, SCARB2, NCAN, SMPD1, MSR1, SIGLEC1, BCAN, LAYN, GDF-8, THY-1, GFR-alpha-1, TNFRSF12A, CPM, CTSC, N2DL-2, IL12, EDA2R, ANGPT1, IL18, FGF-21, KIM1, MMP7, PRSS8, BNP, ACE2, CTSL1, LEP, CA5A, HAOX1, CD33, CDH15, FGFR2, IKZF2, IL3RA, PAEP, RPS6KB1, SFRP1**

#### Mortality

**Social support**
**14 proteins**
**EFNA4, SKR3, TN-R, TNFRSF10A, TNFRSF11A, TRAIL-R2, Gal-9, FGF-23, REN, VSIG2, AMBP, MMP12, ASGR1, PSG1**

**56 proteins**
**UNC5C, VWC2, CLM-6, NBL1, SCARB2, NCAN, SMPD1, MSR1,SIGLEC1, CNTN5, CLEC1B, BCAN, LAYN, GDF-8, WFIKKN1, GFR-alpha-1, NTRK2, SCARF2, TNFRSF12A, GCP5, N2DL-2, CLM-1, SPOCK1, EDA2R, NTRK3, ANGPT1, SLAMF7, PGF, IL-4RA, PAR-1, IL1RL2, IL18, FGF-21, PIgR, SPON2, KIM1, THBS2, CCL17, MMP7, PRSS8, AGRP, MARCO, BNP, ACE2, CTSL1, hOSCAR, TNFRSF13B, CD4,DEFB4A, GGT5, KIR2DL3, LTBP3, NEFL, PLA2G10, SFRP1, SNCG**

**Supplemental Figure 4 Proteins are shared and distinct for the associations between protein signatures with social support (linear regression analysis), and all-cause mortality (Cox regression) and CVD (Cox regression).** The proteins were the significant proteins with FDR adjusted P value less than 0.05 in the minimally adjusted models. The model adjusted for age, sex, education, wealth, and ethnicity.

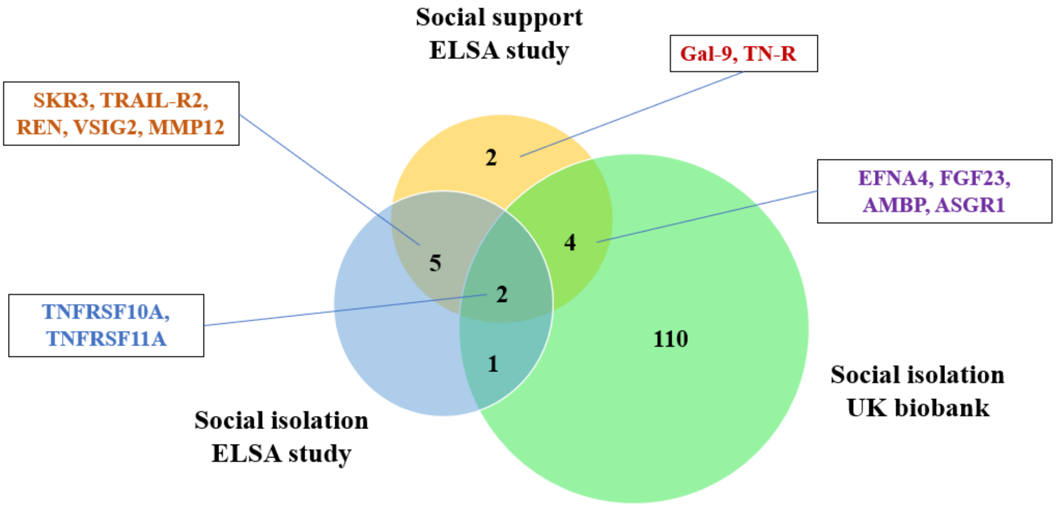


**Supplemental Figure 5 Venn diagram visualizing the overlap of measured protein levels associated with social support after correction and the proteins associated with social isolation shown in the study by Gong, et al. (2025) and that by Shen, et al. (2025).**

The numbers of common and distinct proteins are given, along with the corresponding proteins that are shared with or distinct from those associated with social support.

Shen C, Zhang R, Yu J, Sahakian BJ, Cheng W, Feng J: Plasma proteomic signatures of social isolation and loneliness associated with morbidity and mortality. Nature Human Behaviour 2025, 9(3):569-583.

Gong J, Preminger Z, Steptoe A, Fancourt D: Protein signatures associated with loneliness and social isolation: Plasma proteome analyses in the English Longitudinal Study of Ageing, with causal evidence from Mendelian randomization. Brain Behav Immun 2025, 124:85-94.

**Supplemental Table 1 Associations between social support and proteins**

| Protein | Model 1 | | | | Model 2 | | | |
|---|---|---|---|---|---|---|---|---|
| | Beta | SE | P Value | FDR-adjusted P-value | Beta | SE | P Value | FDR-adjusted P-value |
| EFNA4 | -0.146 | 0.045 | 1.154E-03 | **0.031** | -0.127 | 0.045 | 4.564E-03 | 0.105 |
| SKR3 | -0.169 | 0.044 | 1.161E-04 | **0.002** | -0.127 | 0.043 | 3.317E-03 | 0.101 |
| TN-R | 0.159 | 0.046 | 5.576E-04 | **0.005** | 0.085 | 0.045 | 5.530E-02 | 0.313 |
| TNFRSF10A | -0.174 | 0.044 | 8.402E-05 | **0.002** | -0.156 | 0.044 | 3.682E-04 | **0.020** |
| TNFRSF11A | -0.185 | 0.044 | 3.087E-05 | **0.018** | -0.194 | 0.044 | 1.055E-05 | **0.003** |
| TRAIL-R2 | -0.183 | 0.044 | 2.808E-05 | **0.002** | -0.132 | 0.043 | 1.916E-03 | 0.076 |
| Gal-9 | -0.147 | 0.046 | 1.388E-03 | **<0.001** | -0.140 | 0.046 | 2.207E-03 | 0.076 |
| FGF-23 | -0.203 | 0.046 | 8.080E-06 | **0.025** | -0.182 | 0.045 | 6.409E-05 | **0.004** |
| REN | -0.136 | 0.044 | 1.978E-03 | **0.025** | -0.126 | 0.044 | 3.967E-03 | 0.101 |
| VSIG2 | -0.224 | 0.044 | 4.900E-07 | **0.005** | -0.180 | 0.044 | 4.668E-05 | **0.004** |
| AMBP | -0.151 | 0.045 | 8.680E-04 | **0.031** | -0.150 | 0.045 | 8.591E-04 | **0.040** |
| MMP12 | -0.132 | 0.043 | 2.096E-03 | **0.025** | -0.065 | 0.042 | 1.174E-01 | 0.415 |
| ASGR1 | -0.191 | 0.045 | 2.419E-05 | 0.055 | -0.187 | 0.044 | 2.344E-05 | **0.003** |
| PSG1 | -0.146 | 0.046 | 1.538E-03 | 0.314 | -0.133 | 0.046 | 4.029E-03 | 0.101 |

This table only showed the significant proteins associated with social support in model 1 or model 2 (FDR adjusted P value <0.05).

BMI, body mass index; CVD, cardiovascular disease; CI, confidence interval; HR, hazard ratio.

Model 1: adjusted for age, sex, wealth, education, and ethnicity.

Model 2: additionally adjusted for smoking status, alcohol use, physical activity, and BMI.

**Supplemental Table 2 Associations between positive social support, negative relationship strain and proteins**

| Protein | Model 1 | | | | Model 2 | | | |
|---|---|---|---|---|---|---|---|---|
| | Beta | SE | P Value | FDR-adjusted P-value | Beta | SE | P Value | FDR-adjusted P-value |
| **Positive social support** | | | | | | | | |
| EFNA4 | -0.071 | 0.022 | 1.236E-03 | **0.031** | -0.068 | 0.022 | 1.764E-03 | **0.037** |
| SKR3 | -0.088 | 0.021 | 3.420E-05 | **0.002** | -0.076 | 0.021 | 3.216E-04 | **0.015** |
| TNFRSF10A | -0.084 | 0.021 | 8.696E-05 | **0.005** | -0.086 | 0.021 | 5.467E-05 | **0.005** |
| TNFRSF11A | -0.089 | 0.021 | 3.365E-05 | **0.002** | -0.101 | 0.021 | 2.022E-06 | **0.001** |
| TRAIL-R2 | -0.074 | 0.021 | 4.485E-04 | **0.018** | -0.060 | 0.021 | 4.071E-03 | 0.061 |
| FGF-23 | -0.094 | 0.022 | 2.446E-05 | **0.002** | -0.088 | 0.022 | 7.541E-05 | **0.005** |
| VSIG2 | -0.106 | 0.022 | 1.007E-06 | **<0.001** | -0.088 | 0.021 | 3.976E-05 | **0.005** |
| MMP12 | -0.070 | 0.021 | 8.030E-04 | **0.025** | -0.055 | 0.021 | 1.088E-02 | 0.09 |
| FGFR2 | -0.075 | 0.022 | 7.609E-04 | **0.025** | -0.086 | 0.022 | 1.005E-04 | **0.006** |
| NEFL | -0.075 | 0.019 | 1.028E-04 | **0.005** | -0.064 | 0.019 | 8.071E-04 | **0.028** |
| NXPH1 | -0.073 | 0.023 | 1.357E-03 | **0.031** | -0.074 | 0.023 | 1.271E-03 | **0.032** |
| PSG1 | -0.074 | 0.022 | 9.221E-04 | **0.025** | -0.066 | 0.022 | 3.339E-03 | 0.061 |
| GFR-alpha-1 | -0.063 | 0.021 | 3.000E-03 | 0.055 | -0.066 | 0.021 | 1.513E-03 | **0.035** |
| IL12 | -0.035 | 0.022 | 1.180E-01 | 0.314 | -0.069 | 0.021 | 1.154E-03 | **0.032** |
| Dkk-4 | -0.053 | 0.022 | 1.400E-02 | 0.104 | -0.069 | 0.021 | 1.147E-03 | **0.032** |
| SNCG | -0.051 | 0.022 | 2.400E-02 | 0.133 | -0.069 | 0.022 | 1.942E-03 | **0.038** |
| ASGR1 | -0.064 | 0.022 | 4.000E-03 | 0.057 | -0.075 | 0.022 | 4.773E-04 | **0.019** |
| **Relationship strain** | | | | | | | | |
| LEPR | -0.194 | 0.043 | 7.543E-06 | **0.002** | -0.166 | 0.043 | 1.045E-04 | **0.029** |

This table only showed the significant proteins associated with social support in model 1 or model 2 (FDR adjusted P value <0.05).

BMI, body mass index; CVD, cardiovascular disease; CI, confidence interval; HR, hazard ratio.

Model 1: adjusted for age, sex, wealth, education, and ethnicity.

Model 2: additionally adjusted for smoking status, alcohol use, physical activity, and BMI.

**Supplemental Table 3 Associations between proteins and cardiovascular disease**

| Protein | Model 1 | | | | Model 2 | | | |
|---|---|---|---|---|---|---|---|---|
| | HR | 95% CI | P Value | FDR-adjusted P-value | HR | 95% CI | P Value | FDR-adjusted P-value |
| UNC5C | 1.168 | 1.068,1.276 | 6.700E-04 | **0.008** | 1.147 | 1.049,1.254 | 2.780E-03 | **0.045** |
| CLM-6 | 1.128 | 1.037,1.227 | 5.300E-03 | **0.040** | 1.089 | 1.000,1.185 | 5.020E-02 | 0.254 |
| EFNA4 | 1.159 | 1.063,1.262 | 8.020E-04 | **0.009** | 1.109 | 1.018,1.210 | 1.889E-02 | 0.154 |
| SCARB2 | 1.129 | 1.032,1.235 | 8.614E-03 | **0.049** | 1.104 | 1.008,1.210 | 3.264E-02 | 0.200 |
| NCAN | 0.867 | 0.799,0.940 | 5.775E-04 | **0.008** | 0.917 | 0.843,0.997 | 4.243E-02 | 0.239 |
| SMPD1 | 1.150 | 1.056,1.252 | 1.316E-03 | **0.013** | 1.123 | 1.030,1.224 | 8.820E-03 | 0.101 |
| MSR1 | 1.203 | 1.103,1.312 | 3.39E-05 | **0.001** | 1.172 | 1.075,1.279 | 3.700E-04 | **0.013** |
| SIGLEC1 | 1.230 | 1.129,1.34 | 2.822E-06 | **<0.001** | 1.189 | 1.090,1.296 | 9.661E-05 | **0.004** |
| BCAN | 0.873 | 0.803,0.949 | 1.585E-03 | **0.015** | 0.923 | 0.847,1.007 | 7.122E-02 | 0.277 |
| LAYN | 1.143 | 1.042,1.255 | 4.902E-03 | **0.038** | 1.130 | 1.029,1.240 | 1.069E-02 | 0.118 |
| GDF-8 | 0.883 | 0.812,0.96 | 3.719E-03 | **0.031** | 0.911 | 0.837,0.992 | 3.211E-02 | 0.200 |
| THY-1 | 1.152 | 1.056,1.258 | 1.600E-03 | **0.015** | 1.104 | 1.010,1.207 | 3.026E-02 | 0.199 |
| GFR-alpha-1 | 1.207 | 1.103,1.321 | 4.896E-05 | **0.001** | 1.152 | 1.052,1.262 | 2.345E-03 | **0.040** |
| TNFRSF12A | 1.146 | 1.048,1.253 | 2.947E-03 | **0.026** | 1.121 | 1.024,1.226 | 1.354E-02 | 0.133 |
| SKR3 | 1.224 | 1.12,1.338 | 1.033E-05 | **<0.001** | 1.169 | 1.068,1.279 | 7.629E-04 | **0.021** |
| CPM | 1.159 | 1.067,1.259 | 5.062E-04 | **0.007** | 1.105 | 1.015,1.204 | 2.163E-02 | 0.154 |
| CTSC | 1.152 | 1.063,1.249 | 6.535E-04 | **0.008** | 1.122 | 1.035,1.216 | 5.330E-03 | 0.072 |
| N2DL-2 | 1.127 | 1.034,1.229 | 6.807E-03 | **0.046** | 1.100 | 1.008,1.201 | 3.364E-02 | 0.202 |
| IL12 | 1.117 | 1.029,1.212 | 8.599E-03 | **0.049** | 1.122 | 1.030,1.222 | 8.233E-03 | 0.099 |
| EDA2R | 1.294 | 1.157,1.448 | 7.958E-06 | **<0.001** | 1.243 | 1.111,1.392 | 1.749E-04 | **0.007** |
| ANGPT1 | 0.893 | 0.823,0.968 | 6.125E-03 | **0.043** | 0.905 | 0.835,0.981 | 1.571E-02 | 0.150 |
| TNFRSF10A | 1.263 | 1.158,1.376 | 1.688E-07 | **<0.001** | 1.193 | 1.092,1.302 | 9.767E-05 | **0.004** |
| TNFRSF11A | 1.212 | 1.11,1.323 | 2.031E-05 | **<0.001** | 1.160 | 1.060,1.268 | 1.242E-03 | **0.029** |
| TRAIL-R2 | 1.207 | 1.106,1.317 | 2.929E-05 | **0.001** | 1.124 | 1.027,1.231 | 1.165E-02 | 0.124 |
| IL18 | 1.117 | 1.031,1.211 | 7.102E-03 | **0.046** | 1.087 | 1.002,1.179 | 4.441E-02 | 0.241 |
| FGF-21 | 1.122 | 1.033,1.22 | 6.644E-03 | **0.046** | 1.065 | 0.978,1.161 | 1.488E-01 | 0.450 |
| FGF-23 | 1.190 | 1.093,1.295 | 6.422E-05 | **0.001** | 1.153 | 1.058,1.256 | 1.210E-03 | **0.029** |
| REN | 1.188 | 1.088,1.297 | 1.389E-04 | **0.002** | 1.132 | 1.038,1.236 | 5.460E-03 | 0.072 |
| KIM1 | 1.298 | 1.186,1.42 | 2.176E-08 | **<0.001** | 1.228 | 1.122,1.345 | 1.100E-05 | **0.001** |
| VSIG2 | 1.192 | 1.093,1.299 | 8.079E-05 | **0.001** | 1.147 | 1.051,1.252 | 2.251E-03 | **0.040** |
| AMBP | 1.125 | 1.035,1.223 | 5.981E-03 | **0.043** | 1.076 | 0.988,1.171 | 9.243E-02 | 0.323 |
| MMP7 | 1.217 | 1.116,1.327 | 1.064E-05 | **<0.001** | 1.163 | 1.066,1.270 | 7.715E-04 | **0.021** |
| PRSS8 | 1.210 | 1.11,1.319 | 1.882E-05 | **<0.001** | 1.135 | 1.037,1.242 | 6.265E-03 | 0.079 |
| BNP | 1.283 | 1.173,1.404 | 7.508E-08 | **<0.001** | 1.286 | 1.175,1.407 | 7.000E-08 | **<0.001** |
| MMP12 | 1.278 | 1.166,1.4 | 2.025E-07 | **<0.001** | 1.22 | 1.110,1.341 | 4.453E-05 | **0.003** |
| ACE2 | 1.271 | 1.167,1.383 | 4.645E-08 | **<0.001** | 1.214 | 1.114,1.323 | 1.204E-05 | **0.001** |
| CTSL1 | 1.140 | 1.049,1.238 | 2.198E-03 | **0.020** | 1.135 | 1.044,1.235 | 3.164E-03 | **0.049** |
| LEP | 1.229 | 1.118,1.35 | 2.205E-05 | **<0.001** | 1.120 | 1.000,1.254 | 4.967E-02 | 0.254 |
| CA5A | 1.148 | 1.058,1.244 | 8.942E-04 | **0.010** | 1.102 | 1.015,1.197 | 2.085E-02 | 0.154 |

| | | | | | | | | |
|---|---|---|---|---|---|---|---|---|
| HAOX1 | 1.123 | 1.033,1.222 | 6.938E-03 | **0.046** | 1.088 | 0.999,1.184 | 5.185E-02 | 0.254 |
| ASGR1 | 1.214 | 1.115,1.321 | 8.932E-06 | **<0.001** | 1.135 | 1.039,1.239 | 4.993E-03 | 0.072 |
| CD33 | 1.118 | 1.03,1.213 | 7.608E-03 | **0.047** | 1.105 | 1.018,1.199 | 1.709E-02 | 0.154 |
| CDH15 | 1.131 | 1.033,1.238 | 7.853E-03 | **0.047** | 1.112 | 1.016,1.217 | 2.174E-02 | 0.154 |
| FGFR2 | 1.128 | 1.038,1.226 | 4.678E-03 | **0.037** | 1.081 | 0.994,1.176 | 7.107E-02 | 0.277 |
| IKZF2 | 1.127 | 1.041,1.22 | 3.147E-03 | **0.027** | 1.134 | 1.048,1.228 | 1.961E-03 | **0.040** |
| IL3RA | 1.131 | 1.04,1.231 | 4.168E-03 | **0.034** | 1.105 | 1.016,1.202 | 2.079E-02 | 0.154 |
| PAEP | 1.125 | 1.031,1.227 | 8.092E-03 | **0.048** | 1.111 | 1.018,1.212 | 1.913E-02 | 0.154 |
| RPS6KB1 | 1.144 | 1.055,1.241 | 1.241E-03 | **0.013** | 1.135 | 1.047,1.232 | 2.280E-03 | **0.040** |
| SFRP1 | 1.128 | 1.033,1.233 | 7.511E-03 | **0.047** | 1.091 | 0.999,1.193 | 5.408E-02 | 0.254 |

BMI, body mass index; CVD, cardiovascular disease; CI, confidence interval; HR, hazard ratio.

Model 1: adjusted for age, sex, wealth, education, and ethnicity.

Model 2: additionally adjusted for smoking status, alcohol use, physical activity, and BMI.

#### Supplemental Table 4 Associations between proteins and all-cause mortality

| Protein | Model 1 | | | | Model 2 | | | |
|---|---|---|---|---|---|---|---|---|
| | HR | 95% CI | P Value | FDR-adjusted P-value | HR | 95% CI | P Value | FDR-adjusted P-value |
| UNC5C | 1.104 | 1.026,1.188 | 7.951E-03 | **0.034** | 1.118 | 1.039,1.203 | 2.801E-03 | **0.020** |
| VWC2 | 1.107 | 1.029,1.190 | 6.290E-03 | **0.029** | 1.095 | 1.018,1.177 | 1.433E-02 | 0.069 |
| CLM-6 | 1.102 | 1.028,1.182 | 6.586E-03 | **0.030** | 1.074 | 1.001,1.153 | 4.617E-02 | 0.170 |
| NBL1 | 0.905 | 0.847,0.968 | 3.573E-03 | **0.018** | 0.921 | 0.861,0.985 | 1.695E-02 | 0.079 |
| EFNA4 | 1.174 | 1.094,1.260 | 9.150E-06 | **<0.001** | 1.132 | 1.054,1.215 | 6.538E-04 | **0.007** |
| SCARB2 | 1.142 | 1.062,1.228 | 3.694E-04 | **0.002** | 1.141 | 1.061,1.226 | 3.898E-04 | **0.004** |
| NCAN | 0.850 | 0.794,0.911 | 4.343E-06 | **<0.001** | 0.890 | 0.830,0.954 | 9.868E-04 | **0.010** |
| SMPD1 | 1.167 | 1.085,1.256 | 3.894E-05 | **<0.001** | 1.104 | 1.025,1.189 | 9.185E-03 | **0.046** |
| MSR1 | 1.124 | 1.044,1.211 | 1.902E-03 | **0.010** | 1.123 | 1.043,1.209 | 2.185E-03 | **0.016** |
| SIGLEC1 | 1.167 | 1.086,1.253 | 2.815E-05 | **<0.001** | 1.158 | 1.078,1.245 | 6.512E-05 | **0.001** |
| CNTN5 | 0.869 | 0.809,0.932 | 1.048E-04 | **0.001** | 0.890 | 0.827,0.957 | 1.747E-03 | **0.014** |
| CLEC1B | 0.921 | 0.863,0.982 | 1.236E-02 | **0.049** | 0.912 | 0.855,0.974 | 5.898E-03 | **0.031** |
| BCAN | 0.809 | 0.754,0.869 | 6.817E-09 | **<0.001** | 0.855 | 0.795,0.918 | 2.007E-05 | **<0.001** |
| LAYN | 1.202 | 1.113,1.297 | 2.917E-06 | **<0.001** | 1.174 | 1.087,1.267 | 4.663E-05 | **0.001** |
| GDF-8 | 0.844 | 0.786,0.906 | 3.655E-06 | **<0.001** | 0.902 | 0.839,0.969 | 5.075E-03 | **0.030** |
| WFIKKN1 | 0.869 | 0.809,0.934 | 1.344E-04 | **0.001** | 0.927 | 0.862,0.998 | 4.343E-02 | 0.167 |
| GFR-alpha-1 | 1.187 | 1.100,1.281 | 1.125E-05 | **<0.001** | 1.157 | 1.072,1.249 | 1.844E-04 | **0.002** |
| NTRK2 | 0.901 | 0.840,0.966 | 3.412E-03 | **0.017** | 0.924 | 0.861,0.991 | 2.753E-02 | 0.117 |
| SCARF2 | 1.112 | 1.028,1.204 | 8.627E-03 | **0.037** | 1.114 | 1.029,1.206 | 7.901E-03 | **0.040** |
| TNFRSF12A | 1.175 | 1.091,1.265 | 2.172E-05 | **<0.001** | 1.161 | 1.078,1.251 | 9.278E-05 | **0.001** |
| SKR3 | 1.219 | 1.133,1.312 | 1.472E-07 | **<0.001** | 1.145 | 1.063,1.233 | 3.549E-04 | **0.004** |
| GCP5 | 0.881 | 0.824,0.942 | 2.265E-04 | **0.002** | 0.901 | 0.843,0.964 | 2.531E-03 | **0.018** |
| N2DL-2 | 1.098 | 1.022,1.179 | 1.068E-02 | **0.044** | 1.071 | 0.996,1.151 | 6.420E-02 | 0.211 |
| CLM-1 | 1.104 | 1.031,1.181 | 4.430E-03 | **0.021** | 1.068 | 0.998,1.143 | 5.734E-02 | 0.195 |
| SPOCK1 | 0.884 | 0.826,0.947 | 4.299E-04 | **0.003** | 0.933 | 0.871,1.000 | 4.959E-02 | 0.178 |
| EDA2R | 1.393 | 1.268,1.531 | 1.153E-11 | **<0.001** | 1.362 | 1.239,1.496 | 2.228E-10 | **<0.001** |
| NTRK3 | 0.916 | 0.856,0.980 | 1.118E-02 | **0.045** | 0.949 | 0.886,1.016 | 1.306E-01 | 0.343 |
| TN-R | 0.807 | 0.752,0.866 | 3.787E-09 | **<0.001** | 0.873 | 0.812,0.938 | 2.304E-04 | **0.003** |
| ANGPT1 | 0.903 | 0.846,0.965 | 2.599E-03 | **0.014** | 0.908 | 0.851,0.969 | 3.924E-03 | 0.025 |
| SLAMF7 | 1.158 | 1.082,1.239 | 2.361E-05 | **<0.001** | 1.153 | 1.078,1.233 | 3.720E-05 | **0.001** |
| PGF | 1.201 | 1.115,1.294 | 1.581E-06 | **<0.001** | 1.174 | 1.089,1.266 | 3.118E-05 | **0.001** |
| IL-4RA | 1.144 | 1.067,1.226 | 1.694E-04 | **0.001** | 1.104 | 1.03,1.183 | 5.321E-03 | **0.031** |
| TNFRSF10A | 1.292 | 1.200,1.392 | 1.898E-11 | **<0.001** | 1.231 | 1.142,1.327 | 7.641E-08 | **<0.001** |
| TNFRSF11A | 1.199 | 1.116,1.288 | 9.054E-07 | **<0.001** | 1.183 | 1.101,1.272 | 5.695E-06 | **<0.001** |
| PAR-1 | 1.116 | 1.043,1.193 | 1.430E-03 | **0.008** | 1.099 | 1.028,1.175 | 5.738E-03 | **0.031** |
| TRAIL-R2 | 1.372 | 1.273,1.479 | 5.356E-16 | **<0.001** | 1.262 | 1.169,1.363 | 4.109E-09 | **<0.001** |
| IL1RL2 | 0.895 | 0.834,0.960 | 2.037E-03 | **0.011** | 0.926 | 0.863,0.994 | 3.329E-02 | 0.135 |
| Gal-9 | 1.157 | 1.078,1.242 | 5.665E-05 | **0.001** | 1.120 | 1.043,1.203 | 1.869E-03 | **0.015** |
| IL18 | 1.155 | 1.079,1.236 | 3.760E-05 | **<0.001** | 1.128 | 1.054,1.208 | 5.540E-04 | **0.006** |
| FGF-21 | 1.136 | 1.058,1.221 | 4.783E-04 | **0.003** | 1.091 | 1.015,1.172 | 1.809E-02 | 0.083 |
| PIgR | 1.103 | 1.031,1.181 | 4.400E-03 | **0.021** | 1.042 | 0.974,1.116 | 2.325E-01 | 0.490 |

| | | | | | | | | |
|---|---|---|---|---|---|---|---|---|
| FGF-23 | 1.202 | 1.121,1.289 | 3.130E-07 | **<0.001** | 1.173 | 1.094,1.258 | 8.628E-06 | **<0.001** |
| SPON2 | 1.145 | 1.067,1.228 | 1.787E-04 | **0.001** | 1.128 | 1.051,1.211 | 9.350E-04 | **0.010** |
| REN | 1.207 | 1.121,1.299 | 6.495E-07 | **<0.001** | 1.179 | 1.094,1.269 | 1.592E-05 | **<0.001** |
| KIM1 | 1.397 | 1.298,1.503 | 2.784E-18 | **<0.001** | 1.323 | 1.228,1.425 | 3.668E-13 | **<0.001** |
| THBS2 | 1.119 | 1.044,1.199 | 1.495E-03 | **0.008** | 1.128 | 1.05,1.212 | 1.064E-03 | **0.010** |
| VSIG2 | 1.220 | 1.138,1.308 | 2.725E-08 | **<0.001** | 1.162 | 1.084,1.246 | 2.700E-05 | **0.001** |
| AMBP | 1.135 | 1.059,1.218 | 3.958E-04 | **0.003** | 1.103 | 1.028,1.184 | 6.724E-03 | **0.035** |
| CCL17 | 1.152 | 1.076,1.234 | 5.661E-05 | **0.001** | 1.109 | 1.035,1.187 | 3.293E-03 | **0.023** |
| MMP7 | 1.325 | 1.230,1.427 | 2.842E-13 | **<0.001** | 1.258 | 1.167,1.357 | 3.442E-09 | **<0.001** |
| PRSS8 | 1.275 | 1.185,1.372 | 1.539E-10 | **<0.001** | 1.186 | 1.100,1.279 | 1.107E-05 | **<0.001** |
| AGRP | 1.122 | 1.051,1.198 | 5.870E-04 | **0.004** | 1.102 | 1.032,1.176 | 3.693E-03 | **0.024** |
| MARCO | 1.097 | 1.025,1.173 | 7.426E-03 | **0.033** | 1.117 | 1.043,1.195 | 1.540E-03 | **0.014** |
| BNP | 1.163 | 1.082,1.249 | 4.406E-05 | **<0.001** | 1.148 | 1.068,1.233 | 1.732E-04 | **0.002** |
| MMP12 | 1.376 | 1.274,1.487 | 1.770E-15 | **<0.001** | 1.264 | 1.168,1.368 | 8.982e-09 | **<0.001** |
| ACE2 | 1.157 | 1.075,1.244 | 9.733E-05 | **0.001** | 1.128 | 1.047,1.216 | 1.689E-03 | **0.014** |
| CTSL1 | 1.150 | 1.072,1.232 | 9.349E-05 | **0.001** | 1.145 | 1.068,1.228 | 1.532E-04 | **0.002** |
| hOSCAR | 1.152 | 1.075,1.234 | 6.879E-05 | **0.001** | 1.104 | 1.029,1.184 | 5.736E-03 | **0.031** |
| TNFRSF13B | 1.140 | 1.064,1.222 | 2.193E-04 | **0.002** | 1.103 | 1.029,1.182 | 5.472E-03 | **0.031** |
| CD4 | 1.138 | 1.062,1.220 | 2.650E-04 | **0.002** | 1.104 | 1.031,1.183 | 4.769E-03 | **0.029** |
| ASGR1 | 1.165 | 1.086,1.250 | 2.380E-05 | **<0.001** | 1.121 | 1.043,1.205 | 2.007E-03 | **0.015** |
| DEFB4A | 1.098 | 1.025,1.177 | 7.799E-03 | **0.034** | 1.061 | 0.989,1.137 | 9.854E-02 | 0.283 |
| GGT5 | 1.096 | 1.023,1.174 | 9.546E-03 | **0.040** | 1.090 | 1.017,1.168 | 1.505E-02 | 0.072 |
| KIR2DL3 | 1.121 | 1.048,1.199 | 9.693E-04 | **0.006** | 1.107 | 1.034,1.185 | 3.569E-03 | **0.024** |
| LTBP3 | 1.130 | 1.052,1.215 | 9.006E-04 | **0.005** | 1.125 | 1.046,1.210 | 1.628E-03 | **0.014** |
| NEFL | 1.358 | 1.243,1.483 | 1.914E-11 | **<0.001** | 1.340 | 1.227,1.464 | 1.501e-10 | **<0.001** |
| PLA2G10 | 1.110 | 1.032,1.193 | 5.014E-03 | **0.023** | 1.108 | 1.031,1.191 | 5.569E-03 | **0.031** |
| PSG1 | 1.093 | 1.019,1.172 | 1.259E-02 | **0.050** | 1.079 | 1.006,1.158 | 3.338E-02 | 0.135 |
| SFRP1 | 1.169 | 1.083,1.261 | 6.107E-05 | **0.001** | 1.135 | 1.052,1.225 | 1.090E-03 | **0.010** |
| SNCG | 1.117 | 1.039,1.200 | 2.711E-03 | **0.014** | 1.112 | 1.034,1.196 | 4.405E-03 | **0.028** |

BMI, body mass index; CI, confidence interval; HR, hazard ratio.

Model 1: adjusted for age, sex, wealth, education, and ethnicity.

Model 2: additionally adjusted for smoking status, alcohol use, physical activity, and BMI

**Supplemental Table 5 Association of social support and cardiovascular disease and all-cause mortality**

| | Model 1 | | Model 2 | |
|---|---|---|---|---|
| | HR | 95% CI | HR | 95% CI |
| **CVD** | | | | |
| Social support | 0.723 | 0.588,0.888 | 0.747 | 0.609,0.918 |
| Positive social support | 0.878 | 0.797,0.966 | 0.878 | 0.798,0.967 |
| Relationship strain | 1.047 | 0.868,1.263 | 0.993 | 0.823,1.199 |
| **All-cause mortality** | | | | |
| Social support | 0.654 | 0.548,0.780 | 0.718 | 0.602,0.857 |
| Positive social support | 0.823 | 0.759,0.892 | 0.855 | 0.788,0.927 |
| Relationship strain | 0.955 | 0.807,1.129 | 0.936 | 0.791,1.108 |

BMI, body mass index; CVD, cardiovascular disease; CI, confidence interval; HR, hazard ratio.
Model 1: adjusted for age, sex, wealth, education, and ethnicity.
Model 2: additionally adjusted for smoking status, alcohol use, physical activity, and BMI.

**Supplemental Table 6 Association of protein and social support, cardiovascular disease and all-cause mortality**

| | **Social support** | | | | **CVD** | | | | **All-cause mortality** | | | |
|---|---|---|---|---|---|---|---|---|---|---|---|---|
| Protein | Beta | SE | P Value | FDR-adjusted P-value | HR | 95% CI | P value | FDR-adjusted P-value | HR | 95% CI | P value | FDR-adjusted P-value |
| EFNA4 | -0.146 | 0.045 | 1.154E-03 | **0.032** | 1.159 | 1.063, 1.262 | 8.020E-04 | **<0.001** | 1.107 | 1.029, 1.190 | 9.150E-06 | **0.0289** |
| SKR3 | -0.169 | 0.044 | 1.161E-04 | **0.005** | 1.224 | 1.120, 1.338 | 1.033E-05 | **<0.001** | 1.219 | 1.133, 1.312 | 1.472E-07 | **3.386e-06** |
| TN-R | 0.159 | 0.046 | 5.576E-04 | **0.019** | 0.957 | 0.880, 1.039 | 2.946E-01 | 0.521 | 0.807 | 0.752, 0.866 | 4.282E-10 | **1.161e-07** |
| TNFRSF10A | -0.174 | 0.044 | 8.402E-05 | **0.004** | 1.263 | 1.158, 1.376 | 1.700E-07 | **<0.001** | 1.292 | 1.200, 1.392 | 9.596E-13 | **7.546e-10** |
| TNFRSF11A | -0.185 | 0.044 | 3.087E-05 | **0.002** | 1.212 | 1.11, 1.323 | 2.031E-05 | **<0.001** | 1.199 | 1.116, 1.288 | 9.054E-07 | **1.666e-05** |
| TRAIL-R2 | -0.183 | 0.044 | 2.808E-05 | **0.002** | 1.207 | 1.106, 1.317 | 2.929E-05 | **<0.001** | 1.372 | 1.273, 1.479 | 1.373E-17 | **7.391e-14** |
| Gal-9 | -0.147 | 0.046 | 1.388E-03 | **0.035** | 1.105 | 1.018, 1.199 | 1.754E-02 | 0.0793 | 1.157 | 1.078, 1.242 | 5.665E-05 | **0.0005** |
| FGF-23 | -0.203 | 0.046 | 8.080E-06 | **0.001** | 1.190 | 1.093, 1.295 | 6.422E-05 | **0.0010** | 1.202 | 1.121, 1.289 | 3.130E-07 | **6.646e-06** |
| REN | -0.136 | 0.044 | 1.978E-03 | **0.041** | 1.188 | 1.088, 1.297 | 1.389E-04 | **0.0020** | 1.207 | 1.121, 1.299 | 6.495E-07 | **1.280e-05** |
| VSIG2 | -0.224 | 0.044 | 4.900E-07 | **<0.001** | 1.192 | 1.093, 1.299 | 8.079E-05 | **0.0012** | 1.22 | 1.138, 1.308 | 2.725E-08 | **6.837e-07** |
| AMBP | -0.151 | 0.045 | 8.680E-04 | **0.027** | 1.125 | 1.035, 1.223 | 5.981E-03 | **0.0433** | 1.135 | 1.059, 1.218 | 3.958E-04 | **0.002541** |
| MMP12 | -0.132 | 0.043 | 2.096E-03 | **0.041** | 1.278 | 1.166, 1.400 | 2.000E-07 | **<0.001** | 1.376 | 1.274, 1.487 | 1.770E-15 | **1.628e-13** |
| ASGR1 | -0.191 | 0.045 | 2.419E-05 | **0.002** | 1.214 | 1.115, 1.321 | 8.930E-06 | **0.0002** | 1.165 | 1.086, 1.250 | 2.380E-05 | **0.0002** |
| PSG1 | -0.146 | 0.046 | 1.538E-03 | **0.035** | 1.082 | 0.995, 1.176 | 6.699E-02 | 0.2054 | 1.093 | 1.019, 1.172 | 1.259E-02 | **0.0498** |

This table only showed the significant proteins associated with social support (FDR adjusted P value <0.05), and the association with CVD and all-cause mortality
Model adjusted for age, sex, wealth, education, and ethnicity.

**Supplemental Table 7 Sensitivity analyses using complete case analysis for the association of protein and social support, cardiovascular disease and all-cause mortality**

| | **Social support** | | | | **CVD** | | | | **All-cause mortality** | | | |
|---|---|---|---|---|---|---|---|---|---|---|---|---|
| Protein | Beta | SE | P Value | FDR-adjusted P-value | HR | 95% CI | P value | FDR-adjusted P-value | HR | 95% CI | P value | FDR-adjusted P-value |
| EFNA4 | -0.153 | 0.046 | 8.989E-04 | **0.027** | 1.172 | 1.074,1.28 | 3.803E-04 | **0.005** | 1.17 | 1.089,1.258 | 1.919E-05 | **<0.001** |
| SKR3 | -0.178 | 0.045 | 8.152E-05 | **0.005** | 1.246 | 1.138,1.364 | 1.892E-06 | **<0.001** | 1.218 | 1.129,1.313 | 2.810E-07 | **<0.001** |
| TN-R | 0.173 | 0.047 | 2.289E-04 | **0.008** | 0.948 | 0.872,1.032 | 2.159E-01 | 0.403 | 0.796 | 0.741,0.855 | 4.282E-10 | **<0.001** |
| TNFRSF10A | -0.168 | 0.045 | 2.064E-04 | **0.008** | 1.283 | 1.175,1.401 | 2.937E-08 | **<0.001** | 1.314 | 1.219,1.416 | 9.596E-13 | **<0.001** |
| TNFRSF11A | -0.174 | 0.046 | 1.385E-04 | **0.006** | 1.225 | 1.121,1.339 | 8.254E-06 | **<0.001** | 1.210 | 1.125,1.302 | 3.281E-07 | **<0.001** |
| TRAIL-R2 | -0.174 | 0.045 | 9.429E-05 | **0.005** | 1.212 | 1.108,1.325 | 2.559E-05 | **<0.001** | 1.394 | 1.292,1.505 | 1.373E-17 | **<0.001** |
| FGF-23 | -0.191 | 0.046 | 3.541E-05 | **0.003** | 1.202 | 1.103,1.31 | 2.711E-05 | **<0.001** | 1.204 | 1.121,1.293 | 3.219E-07 | **<0.001** |
| REN | -0.139 | 0.045 | 1.847E-03 | **0.036** | 1.193 | 1.0901.305 | 1.245E-04 | **0.002** | 1.218 | 1.131,1.311 | 1.813E-07 | **<0.001** |
| VSIG2 | -0.233 | 0.045 | 3.181E-07 | **<0.001** | 1.195 | 1.094,1.306 | 8.062E-05 | **0.001** | 1.225 | 1.141,1.315 | 2.130E-08 | **<0.001** |
| AMBP | -0.146 | 0.046 | 1.649E-03 | **0.036** | 1.13 | 1.038,1.231 | 4.812E-03 | **0.030** | 1.15 | 1.071,1.235 | 1.156E-04 | **0.001** |
| MMP12 | -0.137 | 0.044 | 1.754E-03 | **0.036** | 1.286 | 1.173,1.411 | 8.194E-08 | **<0.001** | 1.395 | 1.29,1.509 | 8.436E-17 | **<0.001** |
| ASGR1 | -0.156 | 0.047 | 9.943E-04 | **0.027** | 1.224 | 1.122,1.335 | 5.286E-06 | **<0.001** | 1.173 | 1.091,1.261 | 1.480E-05 | **<0.001** |
| PSG1 | -0.211 | 0.047 | 6.865E-06 | **0.001** | 1.071 | 0.984,1.165 | 1.139E-01 | 0.270 | 1.087 | 1.013,1.166 | 2.109E-02 | 0.077 |
| Gal-9 | -0.134 | 0.047 | 1.444E-03 | 0.072 | 1.100 | 1.011,1.196 | 2.609E-02 | 0.099 | 1.160 | 1.079,1.246 | 5.236E-05 | **<0.001** |

BMI, body mass index; CVD, cardiovascular disease; CI, confidence interval; HR, hazard ratio; SE, Standard Error.
This table only shows the significant proteins associated with social support (FDR adjusted P value <0.05) except for Gal-9 which has been found to be associated with social support in the main analysis.
All models adjusted for age, sex, wealth, education, and ethnicity

**Supplemental Table 8 Sensitivity analyses using complete case analysis for the potential protein mediators in the association of social support and cardiovascular disease**

| Outcome | Total effect | Natural Direct effect | Natural Indirect effect | Proportion mediated, % (95% CI) |
|---|---|---|---|---|
| | HR (95% CI) | HR (95% CI) | HR (95% CI) | |
| **Single mediator** | | | | |
| EFNA4 | 0.732(0.595,0.910) | 0.749(0.607,0.936) | 0.978(0.956,0.993) | 6.2 (1.44, 28.0) |
| SKR3 | 0.731(0.584,0.904) | 0.758(0.612,0.939) | 0.964(0.938,0.983) | 10.0 (3.5, 30.5) |
| TNFRSF10A | 0.750(0.617,0.938) | 0.781(0.644,0.981) | 0.961(0.934,0.981) | 12.2 (5.1, 66.5) |
| TNFRSF11A | 0.750(0.616,0.916) | 0.775(0.641,0.944) | 0.967(0.939,0.986) | 10.3 (3.8, 41.1) |
| TRAIL-R2 | 0.753(0.607,0.906) | 0.777(0.628,0.934) | 0.969(0.947,0.986) | 9.6 (3.2, 32.5) |
| FGF-23 | 0.754(0.606,0.953) | 0.779(0.630,0.975) | 0.968(0.942,0.989) | 10.0 (2.7, 51.8) |
| REN | 0.749(0.601,0.924) | 0.766(0.615,0.951) | 0.978(0.958,0.995) | 6.7 (1.1, 28.6) |
| VSIG2 | 0.746(0.611,0.907) | 0.778(0.628,0.938) | 0.959(0.936,0.982) | 12.5 (3.8, 40.1) |
| AMBP | 0.751(0.634,0.904) | 0.764(0.656,0.925) | 0.982(0.963,0.996) | 5.4 (0.9, 21.1) |
| MMP12 | 0.747(0.628,0.911) | 0.773(0.653,0.941) | 0.967(0.933,0.988) | 10.2 (3.5, 33.4) |
| ASGR1 | 0.719(0.594,0.880) | 0.748(0.619,0.907) | 0.961(0.931,0.985) | 10.5 (3.8, 32.1) |
| **Multiple mediators** | | | | |
| Total mediation | 0.707(0.573,0.887) | 0.782(0.631,0.981) | 0.904(0.854,0.946) | 25.6 (11.0, 83.2) |

All the significant mediators in the single mediation model were put in one model with the multiple mediators.

**Supplemental Table 9 Sensitivity analyses using complete case analysis for exploration of potential protein mediators in the association of social support and all-cause mortality**

| Outcome | Total effect | Natural Direct effect | Natural Indirect effect | Proportion mediated, % (95% CI) |
|---|---|---|---|---|
| | HR (95% CI) | HR (95% CI) | HR (95% CI) | |
| **Single mediator** | | | | |
| EFNA4 | 0.650(0.512,0.776) | 0.665(0.520,0.793) | 0.978(0.955,0.991) | 4.3 (1.4, 11.3) |
| SKR3 | 0.648(0.530,0.783) | 0.670(0.548,0.803) | 0.967(0.945,0.985) | 6.2 (2.3, 14.5) |
| TN-R | 0.661(0.535,0.761) | 0.686(0.555,0.789) | 0.964(0.939,0.983) | 7.3 (2.5, 15.5) |
| TNFRSF10A | 0.651(0.543,0.781) | 0.681(0.570,0.819) | 0.956(0.931,0.981) | 8.5 (3.7, 18.1) |
| TNFRSF11A | 0.652(0.531,0.766) | 0.673(0.551,0.782) | 0.969(0.946,0.986) | 6.1 (1.9, 11.3) |
| TRAIL-R2 | 0.650(0.542,0.797) | 0.687(0.570,0.861) | 0.945(0.915,0.970) | 10.7 (4.9, 27.7) |
| FGF-23 | 0.655(0.536,0.793) | 0.677(0.556,0.817) | 0.967(0.942,0.984) | 6.5 (2.3, 15.2) |
| REN | 0.657(0.532,0.781) | 0.675(0.546,0.807) | 0.974(0.952,0.992) | 5.1 (1.5, 11.4) |
| VSIG2 | 0.650(0.548,0.793) | 0.680(0.578,0.825) | 0.956(0.933,0.978) | 8.5 (3.7, 17.8) |
| AMBP | 0.654(0.530,0.776) | 0.667(0.545,0.796) | 0.981(0.963,0.994) | 3.7 (1.3, 9.5) |
| MMP12 | 0.644(0.535,0.772) | 0.673(0.561,0.809) | 0.956(0.931,0.983) | 8.3 (3.0, 17.0) |
| ASGR1 | 0.653(0.529,0.766) | 0.673(0.546,0.781) | 0.969(0.950,0.988) | 5.9 (2.0,11.2) |
| **Multiple mediators** | | | | |
| **Total mediation** | 0.644(0.506,0.767) | 0.749(0.607,0.894) | 0.86(0.799,0.905) | 29.4 (17.2, 57.2) |

**Supplemental Table 10. Significant functional enrichments in social support-associated proteins**

| Term id | Source | Term name | FDR-adjusted P-value | Intersection size |
|---|---|---|---|---|
| GO:0005035 | GO:MF | death receptor activity | 0.0007 | 2 |
| GO:0046904 | GO:MF | calcium oxalate binding | 0.0208 | 1 |
| GO:0004873 | GO:MF | asialoglycoprotein receptor activity | 0.0208 | 1 |
| GO:0019862 | GO:MF | IgA binding | 0.0234 | 1 |
| GO:0045569 | GO:MF | TRAIL binding | 0.0312 | 1 |
| GO:0019855 | GO:MF | calcium channel inhibitor activity | 0.0403 | 1 |
| GO:0005031 | GO:MF | tumor necrosis factor receptor activity | 0.0403 | 1 |
| GO:0042806 | GO:MF | fucose binding | 0.0403 | 1 |
| GO:0030246 | GO:MF | carbohydrate binding | 0.0403 | 2 |
| GO:0005005 | GO:MF | transmembrane-ephrin receptor activity | 0.0436 | 1 |
| GO:0005159 | GO:MF | insulin-like growth factor receptor binding | 0.0453 | 1 |
| GO:0046875 | GO:MF | ephrin receptor binding | 0.0458 | 1 |
| GO:0004175 | GO:MF | endopeptidase activity | 0.0458 | 2 |
| GO:0005537 | GO:MF | D-mannose binding | 0.0458 | 1 |
| GO:0005003 | GO:MF | ephrin receptor activity | 0.0458 | 1 |
| GO:0004888 | GO:MF | transmembrane signaling receptor activity | 0.0458 | 3 |
| GO:0004190 | GO:MF | aspartic-type endopeptidase activity | 0.0458 | 1 |
| GO:0019865 | GO:MF | immunoglobulin binding | 0.0458 | 1 |
| GO:0070001 | GO:MF | aspartic-type peptidase activity | 0.0458 | 1 |
| GO:0060089 | GO:MF | molecular transducer activity | 0.0481 | 3 |
| GO:0038023 | GO:MF | signaling receptor activity | 0.0481 | 3 |

This table shows the significantly enriched pathways of the 14 proteins associated with social support (FDR-adjusted P-value <0.05).
